# Analysis of Multiple Myeloma Drug Efficacy

**DOI:** 10.1101/2024.08.03.24311450

**Authors:** Alexandre Matov

## Abstract

**Introduction:** Multiple myeloma (MM) is an incurable plasma cell neoplasm. MM-specific alterations in methylation status cause gradual epigenetic changes and lead to pre-MM disease states, such as Monoclonal Gammopathy of Undetermined Significance (MGUS) and Smoldering MM (SMM). The communication between MM cells and the bone marrow (BM) stromal cells serves a pivotal role in MM development by supporting transformed cell growth and proliferation. MM cells are known to modify the BM microenvironment through secretion of exosomes, which enhances disease progression by the induction of angiogenesis, immune suppression as well as drug resistance. This form of intercellular communication is thought to be mediated by several types of cargo molecules prevalent in exosomes, including microRNAs (miRs).

**Methods:** The main obstacle in the treatment of MM is the difficulty in eliminating the residual cancer cells. Even if there are multiple treatment options, none is curative, and remissions have an unpredictable relapse onset. We attempt to address the two hurdles in terms of the difficulty in predicting the duration of remission and the challenge, which currently remains out of reach, treatment regiments that guarantee cancer-free bone marrow and propose a computational strategy based on our analysis of patient samples and patient cultures.

**Results:** Our method will allow performing quantitative live-cell companion diagnostics by evaluating the relative contribution of different signaling pathways in drug resistance and response via quantitative exosome imaging, beyond MM, in primary tumor cells originating from different organs and tissues.

**Conclusions:** Our approach will allow us to identify putative drug targets for the treatment of refractory disease for which currently there is no known suitable treatment regimen in acute myeloid leukemia, primary pancreatic, and bone metastatic prostate tumors.

## INTRODUCTION

### Available Drugs in MM

MM (Palumbo and Anderson, 2011) remains incurable – a fact underscored by a large number of drugs (available as 37 combinations and counting) used to treat the disease. The available drugs belong to categories chemotherapy (Child et al., 2003), steroids (McIntyre, 1979), immunotherapy (Cohen et al., 2025; Garfall et al., 2015), proteasome inhibitors (Anderson, 2009), histone deacetylase inhibitors (Hideshima et al., 2016; Prince et al., 2008), and monoclonal antibodies (Raje and Longo, 2015), and drugs from different categories (Laubach, 2019) are used together as combination therapies. Dexamethasone, for instance, is a corticosteroid medication offered for the treatment of relapsed/refractory MM in combinations with different classes of drugs like isatuximab, plitidepsin, selinexor, carfilzomib, daratumumab, lenalidomide, and bortezomib, which have different mechanisms of action (Attal et al., 2017; Chari et al., 2019; Facon et al., 2024; Facon et al., 2019; Landgren et al., 2019; Spicka et al., 2019). The complexity of the MM regimens (Choudhry et al., 2018) highlights the incredible ability of the malignant plasma cells to survive and suggests the need to develop quantitative methods for evaluation of both the short and long-term effects of drug treatments on their cellular target before initiating treatment.

### Drug Resistance and Residual Disease

The main hurdle in treatment of MM is that even if there are multiple treatment options, none is curative and remissions have an unpredictable relapse onset. With this contribution we will attempt to address the difficulty in anticipating drug resistance based on *ex vivo* treatment analyses of patient-derived primary cells. Our model system will utilize cells extracted from bone marrow biopsies and cultured in microfluidics to mimic the tumor microenvironment. We will utilize quantitative dual-color high resolution time-lapse live-cell confocal microscopy to image the motion of patient exosomes as well as the changes in morphology of the histone acetylation. Our central hypothesis is that rates of multiple myeloma cells exosome secretion can be correlated with drug response and, thus, predict resistance to treatment. We measure the associated with drug treatment changes in the rates of exosome generation as the number of new particle tracks as well as the motion of the exosomes, i.e., their speeds of translocation on the cell surface during secretion for instance before and after treatment with Daratumumab (Facon et al., 2019) and/or vincristine (Wang et al., 2023) as well as bortezomib (Attal et al., 2017) and/or MPTOG413 (Huang et al., 2019) to investigate the mechanisms behind drug resistance (see schematics on Fig. 1).

**Figure 1.**
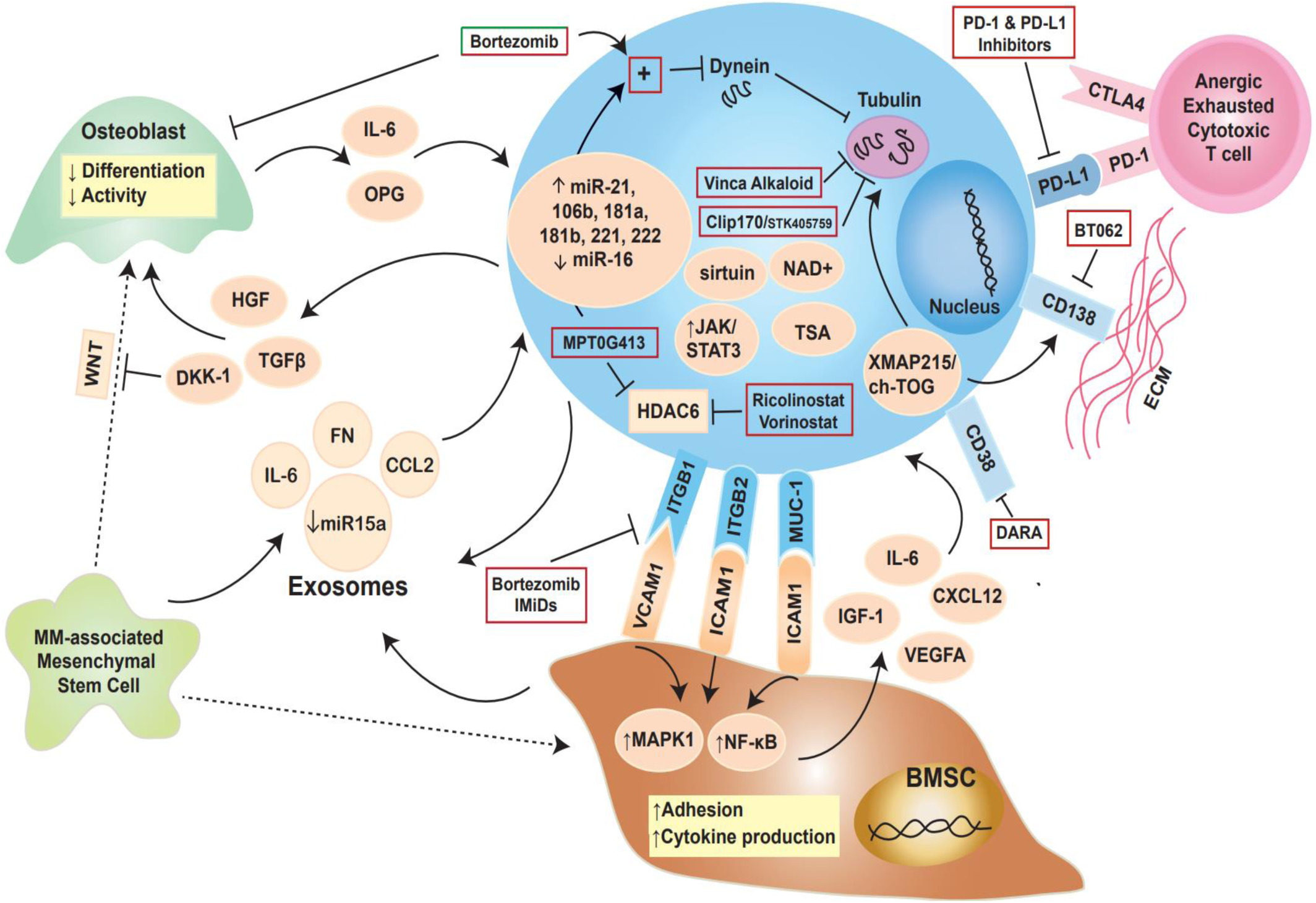
Regulation of drug resistance in MM. The myeloma cell is in blue and the interplay with cellular and other components, in particular those related to microtubule (MT) regulation, is depicted on the schematics. Secretion of exosomes allows the myeloma cell to communicate with the tumor microenvironment. A panel of miRs has been implicated in pathogenesis and myeloma cell proliferation (Pichiorri et al., 2008; Roccaro et al., 2009). Daratumumab is an anti-CD38 monoclonal antibody, which prevents SMM from progressing to MM (Dimopoulos et al., 2024). Histone deacetylase 6 (HDAC6) binds polyubiquitinated proteins and dynein motors and transports this protein cargo to the aggresome for degradation. Combination therapy can inhibit adherence of MM cells to bone marrow stromal cells (BMSC) and reduce VEGF and IL-6 levels and myeloma cell growth (Huang et al., 2019). STK405759, a member of the furan metotica family, as a novel, potential tubulin inhibitor for MM treatment (Rozic et al., 2015). The MT polymerase chTOG is implicated in tumorigenesis (Lauffart et al., 2002). Exosomes regulate MM-related processes including osteolysis, angiogenesis, immune suppression, and drug resistance (Menu and Vanderkerken, 2022). The MT-depolymerizing drug vincristine is used in combination therapy to treat MM (Wang et al., 2023).

### Role of Exosomes

Exosomes are critical to the progression of cancer through the transport of a variety of molecular cargos, including proteins, lipids, and nucleic acids. MM cell-derived exosomes promote BM stromal cells viability, induce changes in pro-survival pathways, and the inhibition of their uptake suppresses the functional response in BM stromal cells (Zheng et al., 2019). Cellular exosome uptake depends on cholesterol-rich membrane micro-domains called lipid rafts (Plebanek et al., 2015). Despite recent advances in treatment, MM remains an incurable malignancy, and *in vitro*, *ex vivo* and *in vivo* approaches have identified lipid rafts to constitute a new target in MM (Mollinedo et al., 2010). Novel compounds target and accumulate in MM cell membrane rafts, inducing apoptosis through the co-clustering of rafts and death receptors. Raft disruption by cholesterol depletion inhibited drug uptake by tumor cells as well as cell killing (Plebanek et al., 2015). Modulation of cholesterol flux can, therefore, lead to potent inhibition of tumor cell-derived exosomes, which could deliver a marker for *ex vivo* testing of drug uptake. Exosomes produced by MM cells have been shown to promote angiogenesis and immunosuppression, both crucial events in MM progression (Wang et al., 2014). Microfluidics-based platforms (Aleman et al., 2019; Zhang et al., 2014) are utilized for *ex vivo* culturing of primary MM cells to emulate the dynamic physiology of the bone niche.

### Histone Acetylation and Role of miRs

Epigenetic nuclear reorganization and changes in the image texture of histone markers and morphology (Farhy et al., 2019) after acetylation can serve for the purposes of pre-clinical microscopic evaluation of drug efficacy (Matov, 2024f). Our objective is to perform such analyses in patient-derived cells and validation the analysis based on actual patient response in the clinic. Further, we will be performing longitudinal analysis of body fluids with the objective of monitoring gradual changes (Matov, 2024h) in the regulation of the BM. To this end, we will monitor changes in the abundance levels of nucleic acids such as miRs. In this context, we hypothesize that drug resistance in MM could be overcome by sensitizing tumor cells with small molecules that correct the dysregulated miRs associated with refractory disease. As pre-cancerous changes in methylation status have been linked to changes in the miRs expression levels (Saviana et al., 2023; Suzuki et al., 2012), we will investigate the pre-myeloma epigenetic changes to test whether they can be reversed also by targeting the dysregulated miRs.

### Drug Selection

One avenue to address the difficulty in anticipating drug resistance in MM is the *ex vivo* drug treatment analyses of patient-derived primary cells imaged by quantitative multi-color high resolution time-lapse live-cell confocal microscopy to track the motion (Matov, 2024f; Matov, 2024g) of exosomes as well as the changes in histone morphology after acetylation. The rates of MM cells exosome secretion are regulated by a variety of genes and changes in the rates of secretion can be correlated with drug response and, thus, predict resistance to therapy. Our hypothesis is that differential regulation affects the associated with drug treatment changes in the rates of exosome secretion, such as the (i) number of new exosome particle tracks as well as the (ii) speed of their motion on the cell surface membrane during secretion before and after drug treatment. As the drug response motion trajectory differences between exosomes will be reflective of the cell type, the approach described in this article will offer a mechanistic insight that will contribute to understanding the variety in tumor cell response.

Currently, most patients receive very similar upfront regimens regardless of genotype or phenotype. Very little, if any therapy, is genome-guided, except perhaps for the intensity of upfront therapy as a function of prognostic markers. The genetic analysis process does not involve a microscopic consideration of drug-target engagement at the cellular level. There are multiple factors that pre-determine the ability of a particular drug to engage its target effectively. Important is the ability to precisely measure and provide statistical analysis of the intercellular communication between primary tumor cells under drug treatment in a lab setting. Even if initial treatments induce a state of remission, the disease eventually returns (Lonial, 2010) and, in most cases, does respond to other therapies, even though resistant to the previously given one. This type of drug response indicates that it is important to develop tools for an upfront optimal drug selection from the presently available treatment options at the onset of therapy.

## RESULTS

### Changes in Urinary Nucleic Acids

We performed longitudinal analysis of body fluids with the objective of monitoring gradual changes (Fig. 2) in the regulation of the bone marrow expressed in alteration in the abundance levels of nucleic acids such as miRs (Pichiorri et al., 2008; Roccaro et al., 2009). Our analysis of established biomarkers for MM indicated changes in the levels of two biomarkers, miR-181a-5p (maximal abundance level we detected in a healthy individual was 1,899) and miR-221-5p (maximal abundance level we detected in a healthy individual was 184), toward a disease state. Longitudinal analysis of nucleic acids will allow for the development of novel targeted drugs, such as the liver-specific miR-122 inhibitor miravirsen (Gebert et al., 2014). Recent literature has described a plethora of examples of miRs (Matov, 2024h) that could be targeted in disease. Therefore, monitoring of their levels in healthy individuals (Matov, 2024h) and patients undergoing disease treatment will likely provide valuable datasets for the pharmaceutical industry as well as for practicing physicians, ultimately allowing them to select the most efficacious treatment sequence and drug combinations for each patient. Our current focus is the identification of personalized biomarker panels whose measurable changes, exceeding the intrinsic stochastic variability in the post-transcriptional regulation of gene expression in normal physiology, uniquely define the degenerative diseases at an early stage when better treatment options are available.

**Figure 2.**
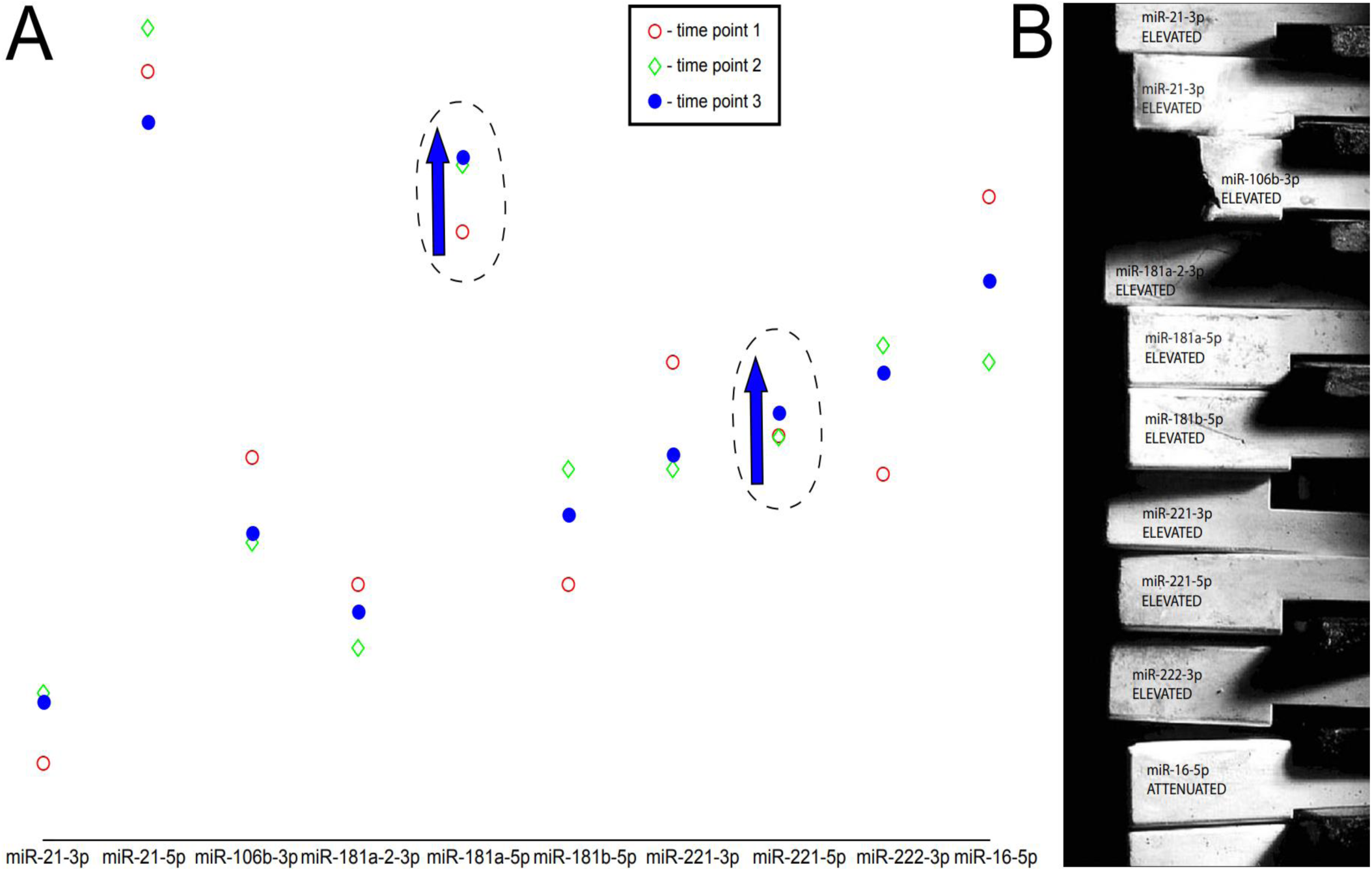
An established panel of miRs deregulated in multiple myeloma, which are also identified in our preliminary urine data. (A) The miRs are listed on the X-axis and log_2_ of their abundance levels are on the Y-axis. See the color-coding figure legend for time progression labeling: red circles show the abundance levels in time point 1, green rhombs show the abundance levels in time point 2, and blue circles show the abundance levels in time point 3. Based on the list of established biomarkers for multiple myeloma, we show the changes occurring for one person over three time points taken at two months intervals. All biomarkers but the last one are upregulated in multiple myeloma (see (B) and (Pichiorri et al., 2008; Roccaro et al., 2009)); therefore, for this particular subject, two biomarkers, miR-181a-5p (maximal abundance level we detected in a healthy individual was 1,899) and miR-221-5p (maximal abundance level we detected in a healthy individual was 184) - both encircled on the figure - albeit minimally, are changing toward a disease state. miR-21-3p is elevated in the second time point, but is then lowered in the third. (B) Schematic representation depicting the dysregulation of miRs in multiple myeloma from (A); therapeutic strategies can be designed to apply drug treatments that restore the miRs’ abundance levels back toward correcting the expression of their underlying gene toward regulation as in normal physiology. Longitudinal analysis of urine samples can, thus, provide a non-invasive drug efficacy readout assay.

Our aim is to outline a rationale for overcoming drug resistance in MM by sensitizing tumor cells with small molecules that correct the dysregulated miRs, which have been associated with refractory disease. As pre-cancerous changes in methylation status have been linked to changes in the miRs abundance levels (Saviana et al., 2023), one could also investigate the pre-myeloma epigenetic changes to test whether they can be reversed also by targeting the dysregulated miRs. Elucidating the specific molecular mechanisms of drug resistance will allow for a better understanding of the reasons behind the failure of all existing treatment regimens to cure MM. In this context, the utilization of a real-time computer vision system (Matov, 2024g) that can reliably evaluate drug action *ex vivo* will also provide an additional platform for functional testing of putative compounds and combination therapies.

### Analysis of Multiple Myeloma Exosomes

#### Real-Time Quantitative Imaging in 2D

We imaged exosomes in MM1S cells in 3D using spinning disc confocal microscopy (Fig. 3). To track the motion of exosomes, we will pre-process images making up the microscopy video and select features (Shi and Tomasi, 1994) for motion tracking on-the-fly. Using this data, we will define exosomes’ dynamics signatures to distinguish between drug resistance signaling pathways. For each exosome type, based on different exosome markers, we can extract a set of distinctive key features (Rublee et al., 2011), identify descriptor vectors of the features and compute motion metrics to distinguish between drug-resistant and sensitive profiles in cells derived from patients’ BM, and elucidate mechanisms of resistance. After classifying all exosome motion trajectories as sensitive or resistant, within the resistant trajectories, we will assign different classes of motion behavior to differentiate between the mechanisms of drug resistance. Our analysis will allow us to identify the relative contributions of different types of exosomes to different resistance mechanisms and pathways. As the differences between exosomes are reflective of the underlying exosome cell type, our novel research tool will offer a mechanistic insight that will contribute to understanding the variety in tumor cell response.

**Figure 3.**
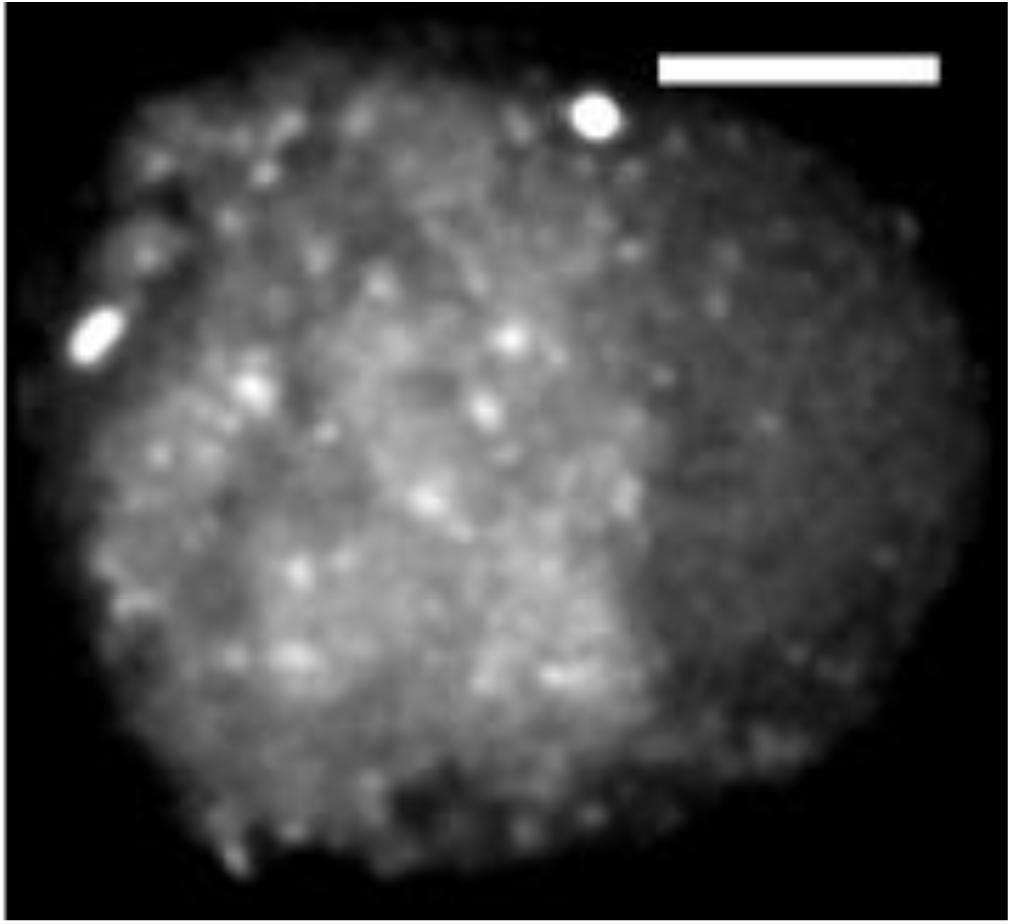
Exosomes in a multiple myeloma cell. Spinning-disk confocal image of the suspension MM1S cell lines acquired five days after labeling with CD63-RFP. A grayscale maximal intensity projection image was aggregated from 20 z-sections to display the 3D volume of exosomes in 2D. The image has been processed with a low pass filter with a cut-off (σ = 1.25) corresponding to the optical limit of the collection frequencies based on the diffraction limit of a 1.45 NA objective. For visualization purposes, the low intensity values were computationally clipped. Scale bar equals 5 µm.

This analysis will, therefore, allow for the identification of the key signaling pathways involved in resistance, which may contribute to the personalization of drug regimens and the functional testing of putative compounds. This computational tool will allow supplementing existing genetic tools with an orthogonal approach to elucidating drug resistance in quantitative detail based on the phenotypical changes in the intracellular communications between tumor cells and their immediate microenvironment. A key novelty is that our computational platform outputs results in real-time during image acquisition (Matov, 2024f), which will facilitate and speed up research efforts by providing instant delivery of data and statistical analysis.

#### Real-Time Quantitative Imaging in 3D

We generated preliminary imaging data (Fig. 4) using 3D spinning disk confocal microscopy with the glucocorticoid-sensitive cell line MM1S (Greenstein et al., 2003) without a drug and after a 24-hour treatment with dexamethasone at concentrations of 60 nM, 120 nM, and 250 nM (the lab IC50 value is 300 nM (Matulis et al., 2016)). To obtain preliminary data, we performed overnight lentiviral transduction and were able to observe the expression of the fluorescent tetraspanin exosome marker CD63-RFP (enriched in extracellular proteins) (Lotvall et al., 2014) 16 hours post infection, which suggests that we could deliver a phenotypical evaluation of the drug-treatment regimen that overcomes the drug resistance mechanisms in refractory disease within a week of patient biopsy collection.

**Figure 4.**
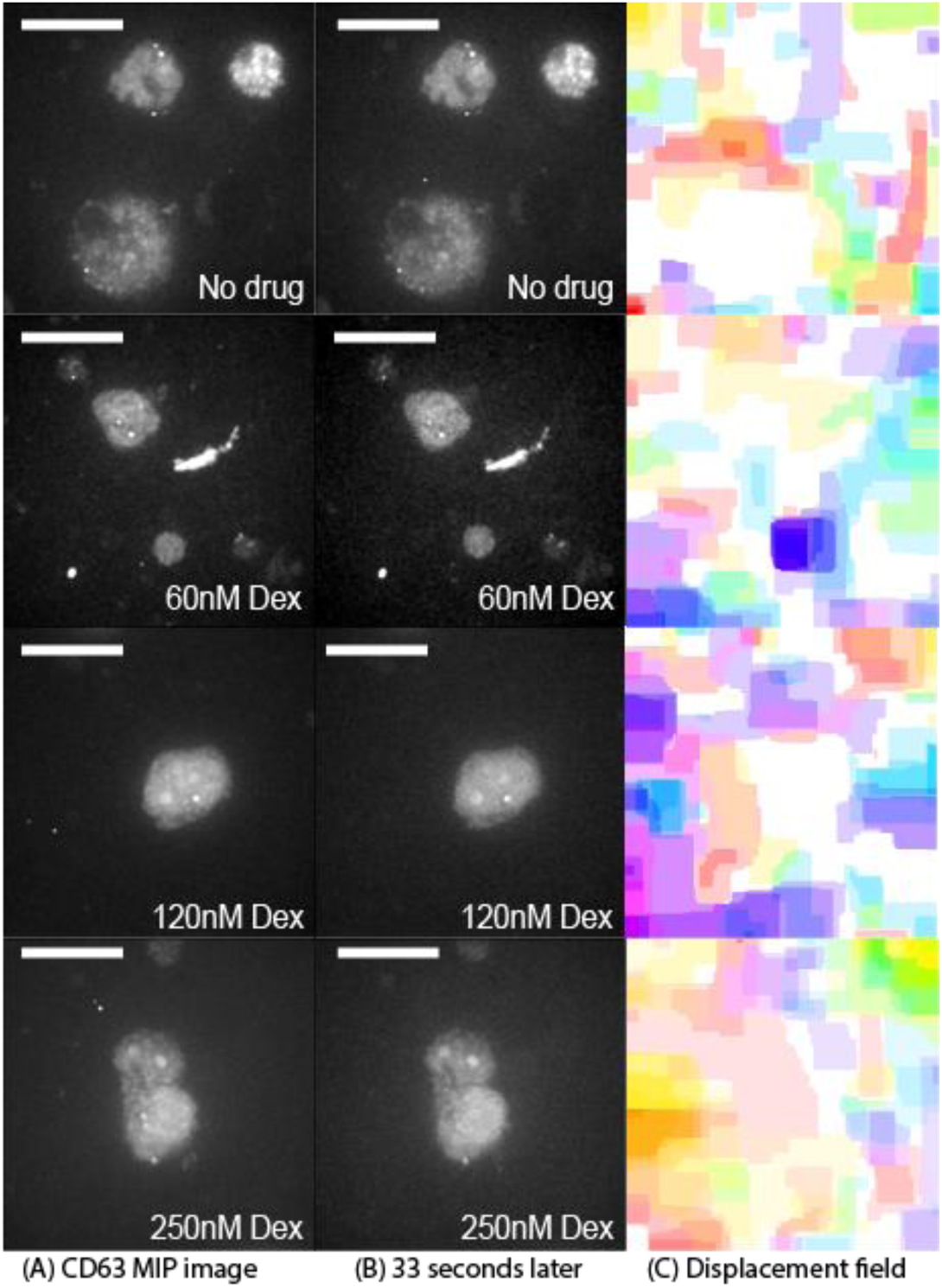
3D SIFT displacement field analysis of exosome images of untreated cells and of cells 24 hours after treatment with 60 nM, 120 nM or 250 nM dexamethasone. Spinning-disk confocal images of cells from the suspension MM1S cell line at high resolution. Images were acquired 11 days after labeling with CD63-RFP via lentiviral transduction, and for some cells, was performed a 24-hour titration with dexamethasone at concentrations 60 nM, 120 nM, and 250 nM. 3D z-stacks, consisting of 20 2D slices with a 500 ms exposure time at a 2 µm vertical spacing, were acquired. The lateral resolution of the objective is 220 nM. The temporal frequency with which the 3D image stacks were obtained was 11 seconds. Maximal intensity projection (MIP) images were generated in which every pixel has the highest intensity value from the z-axis vector for each x-y position. This transformation allows for the visualization of the combined 3D volume of exosomes in a 2D figure. For evaluation of the 3D SIFT Flow and displacement fields, adjacent in time 2D slices were combined in single 3D files/triplets. Thus, the displacement computation between (A-B) is done over a period of 33 seconds. The cells were immobilized with poly-l-ornithine for imaging. For this reason, a vast proportion of the SIFT flow measured and displayed in (C) is due to the diffusion of culture medium. To visualize the flow fields in (C), each pixel denotes a flow vector for which the angular orientation and magnitude value are represented by the hue and the saturation of the pixel, respectively. Scale bar equals 10 µm.

To compute a faithful representation of the image stack within the entire time-lapse sequence, we will utilize scale-invariant feature transform (SIFT) (Lowe, 1999a; Lowe, 1999b), which is an object recognition system has been developed that uses local image features that are invariant to image scaling, translation, and rotation, and partially invariant to illumination changes and affine or 3D projection. The features in SIFT share similar properties with neurons in the inferior temporal cortex that are used for object recognition in our vision. The features are efficiently detected through a staged filtering approach that identifies stable points in scale space (Lindeberg, 1993; Lindeberg, 1998). We performed SIFT in 3D (Scovanner et al., 2007) and Fig. 4 shows our preliminary results for computing SIFT 3D Flow (Liu et al., 2011), which allowed us to compute pixel displacement fields in these time-lapse image series as a first estimation of exosome motion.

To assign a foreground / background label for each pixel in the time-lapse sequences, we will utilize real-time 3D image segmentation algorithms. To simultaneously optimize both the segmentation and assignment steps, we will utilize a 3D CNN (Hou et al., 2019) using separable convolution with dilation, which is suitable for real-time analysis. Adding dilation rates will allow us to capture multi-scale feature representations without significantly increasing the computation cost of 3D convolution. Because of the low signal-to-noise ratio in the high-resolution exosome images due to photobleaching, we expect false-negative selections during the image segmentation phase. In this context, we will utilize a Bayesian algorithm, which addresses the problem of decision-making given incomplete information. Incomplete information could arise from datasets where certain features are missing in various locations. Importantly, through Bayesian methods, decision-making with uncertainty estimates can be produced (Chandra et al., 2019). Further, as an improvement to spinning disc confocal microscopy, we will perform 3D imaging using lattice light-sheet microscopy (Matov, 2024e) to image intercellular communication via exosomes to ensure a minimal damage to the cells and long-term imaging without photobleaching at comparable x-y-z resolution limits (Chen et al., 2014).

### Analysis of Patient Circulating Tumor Cells

Changes in the number of CTCs serve as a prognostic and predictive marker in the management of metastatic disease (Sung et al., 2013; Sung et al., 2012; Tasaki et al., 2014). Changes in morphology and subcellular localization of proteins of interest can also be utilized to evaluate disease progression and drug efficacy. We developed an automated platform (Matov, 2024f) for the measurement of MT bundling patterns and drug-induced inhibition of nuclear accumulation of the androgen receptor (AR) in circulating tumor cells (CTCs), which can be used to predict the clinical response to drug treatment. Our computer vision algorithm performs identification of CTCs (Matov, 2024c) in low magnification confocal microscopy images of peripheral blood of metastatic prostate cancer (PC) patients stained with cell surface makers on coverslips (Matov, 2024b). Furthermore, in high magnification images we evaluate changes in the morphology of the MT cytoskeleton (Matov, 2024e) and compute the sub-cellular localization of AR and after drug treatment. We present new results on the quantification of the percentage of nuclear AR of the volume of the cell, as readouts of effective drug-target engagement in patient-derived CTCs (Fig. 5).

**Figure 5.**
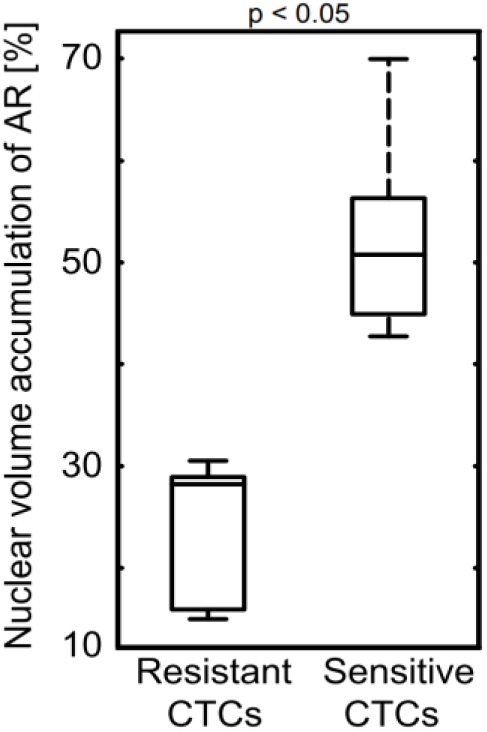
AR nuclear accumulation based on 3D analysis of 35 2D stacks of CTCs from PC metastatic patients. Of the 10 coverslips with patient samples we analyzed, half were sensitive and half were resistant to clinical treatment with docetaxel. The drug sensitive patients had an average of 52% of nuclear accumulation of AR. The drug resistant patients had an average of 24% of nuclear accumulation of AR. Images were acquired at 63x magnification.

### Analysis of Patient Organoids and Metastatic Potential

We derived rectal cancer (RC) organoids from resected tissue of a metastatic rectal tumor protruding from the sternum. When the organoids exceeded size of 150 µm in diameter, they began forming intestinal glands or crypts (Lieberkühn, 1782). The intestinal epithelium is an integral component of innate immunity. The intestinal epithelial cells shape multitubular invaginations that form crypts, which increase the absorption surface of the tissue. At the base of the crypts, the intestinal stem cell niche enables the constant regeneration of the intestinal lining (e.g., enterocytes, endocrines, or goblet cells). These cells can proliferate, differentiate and move upwards, where they can be replaced after a couple of days. In our organoid culture, the patient cancer cells readily formed organoids larger than ½ mm in diameter with some of the crypts exceeding 100 µm in diameter (Fig. 6). To evaluate the metastic potential of the organiods, we used a cell migration transwell asssay (Chen, 2005) and observed organoid cells protruding overnight through the insert membrane (Vid. 1), which indicated a strong metastatic potential of the tumor.

**Figure 6.**
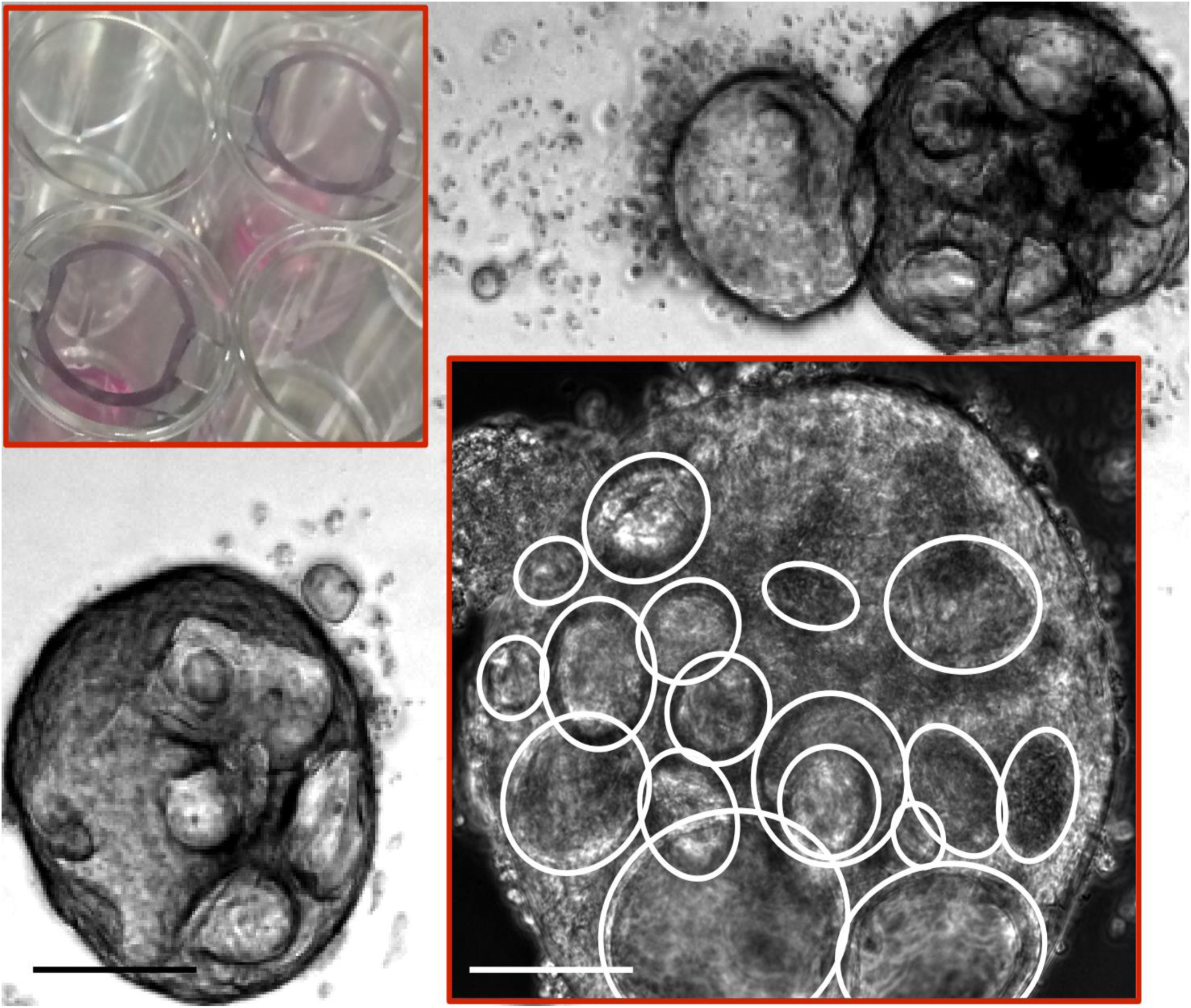
Sternum metastasis RC organoids. The organoids exhibit strong metastatic potential assessed via a transwell assay – see upper inset and Vid. 1. Crypts are clearly visible in large organoids (see white ovals on the lower inset). Scale bars equal 100 µm. Transmitted light microscopy, 4x magnification. Inset: Phase contrast microscopy, 20x magnification.

### Analysis of Ferroptosis and Elimination of Residual Disease

#### Drug-Tolerant Persister Cells

Ferroptosis has a dual role in cancer. It plays a role in tumor initiation, tumorigenesis, which depends on inflammation-associated immunosuppression triggered by ferroptotic damage (Chen et al., 2021) and later, during treatment, in tumor suppression (Hangauer et al., 2017). We sought to identify therapeutically exploitable vulnerabilities in cancer cells. Initial data was obtained with a metastatic RC organoid (we established from 70 mg resected lung tissue containing metastatic tumor protruding from the sternum) with the standard first line metastatic colorectal cancer chemotherapy FOLFOX. We plated organoids with diameters 40-80 µm in 24 wells and performed titration with concentration of 1 µM (Fig. 7), 10 µM, and 100 µM for two weeks. We repeated the same with docetaxel with concentration of 100 nM and 1 µM (Fig. 8), and the experiment showed clear residual disease measured by CellTiter-Glo assay (ATP luminescence had values of up to 6,000). This was the first example of “persister” cancer cells in 3D patient-derived organoids cultured in 30 µL Matrigel drops (van de Wetering et al., 2015), i.e., in their natural 3D environment and not as single patient cells cultured on a flat plastic surface in a 2D Petri dish.

**Figure 7.**
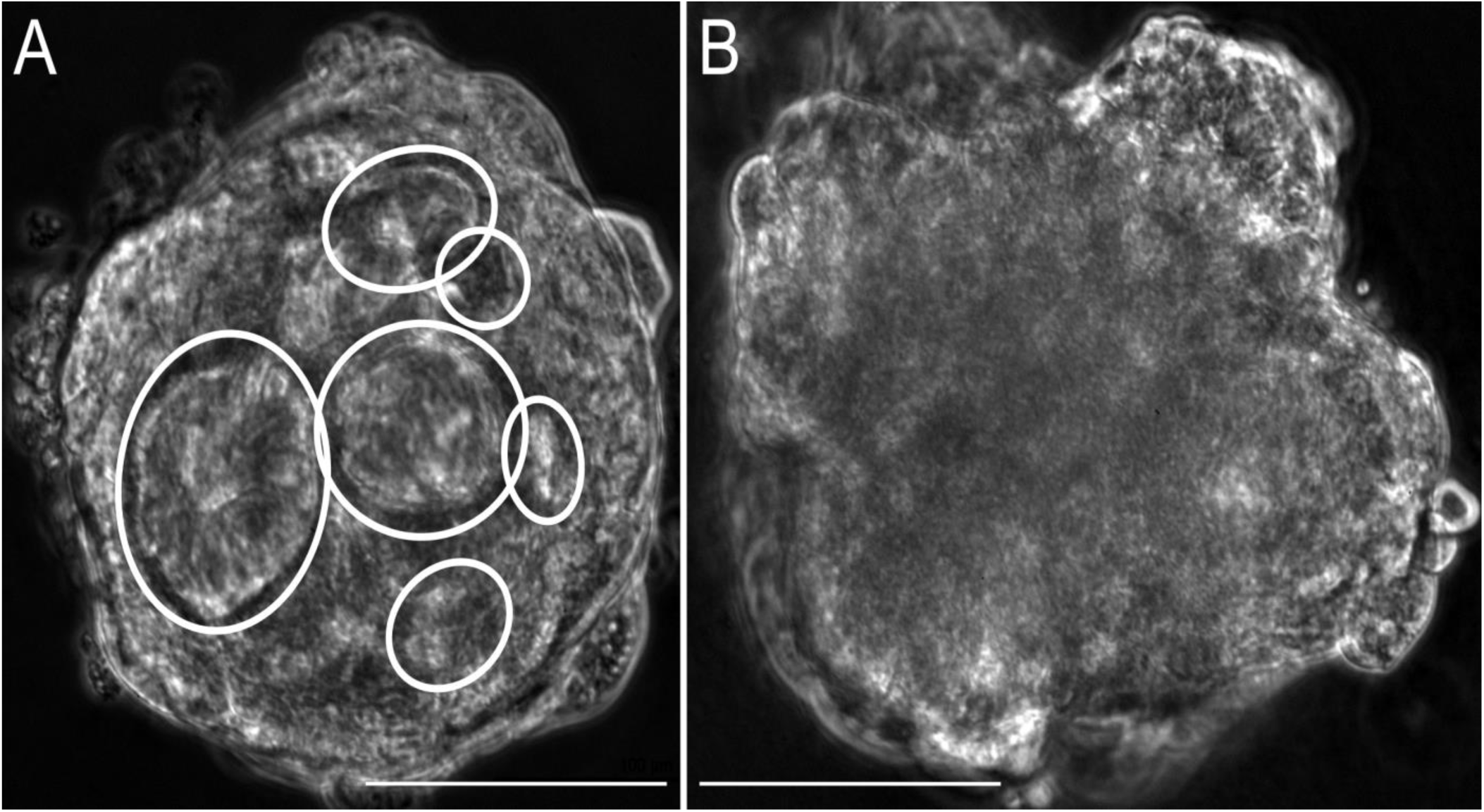
Sternum metastasis RC organoids receding in diameter after treatment with FOLFOX at 1 µM. (A) Crypts are clearly visible on this image and even if the organoid edges are still intact, they appear very uneven after 48 hours of FOLFOX treatment. (B) After additional 13 days of drug treatment, i.e., 15 days in total, the crypts are no longer visible, but the organoids did not receed. Scale bars equal 100 µm. Phase contrast microscopy, 20x magnification.

**Figure 8.**
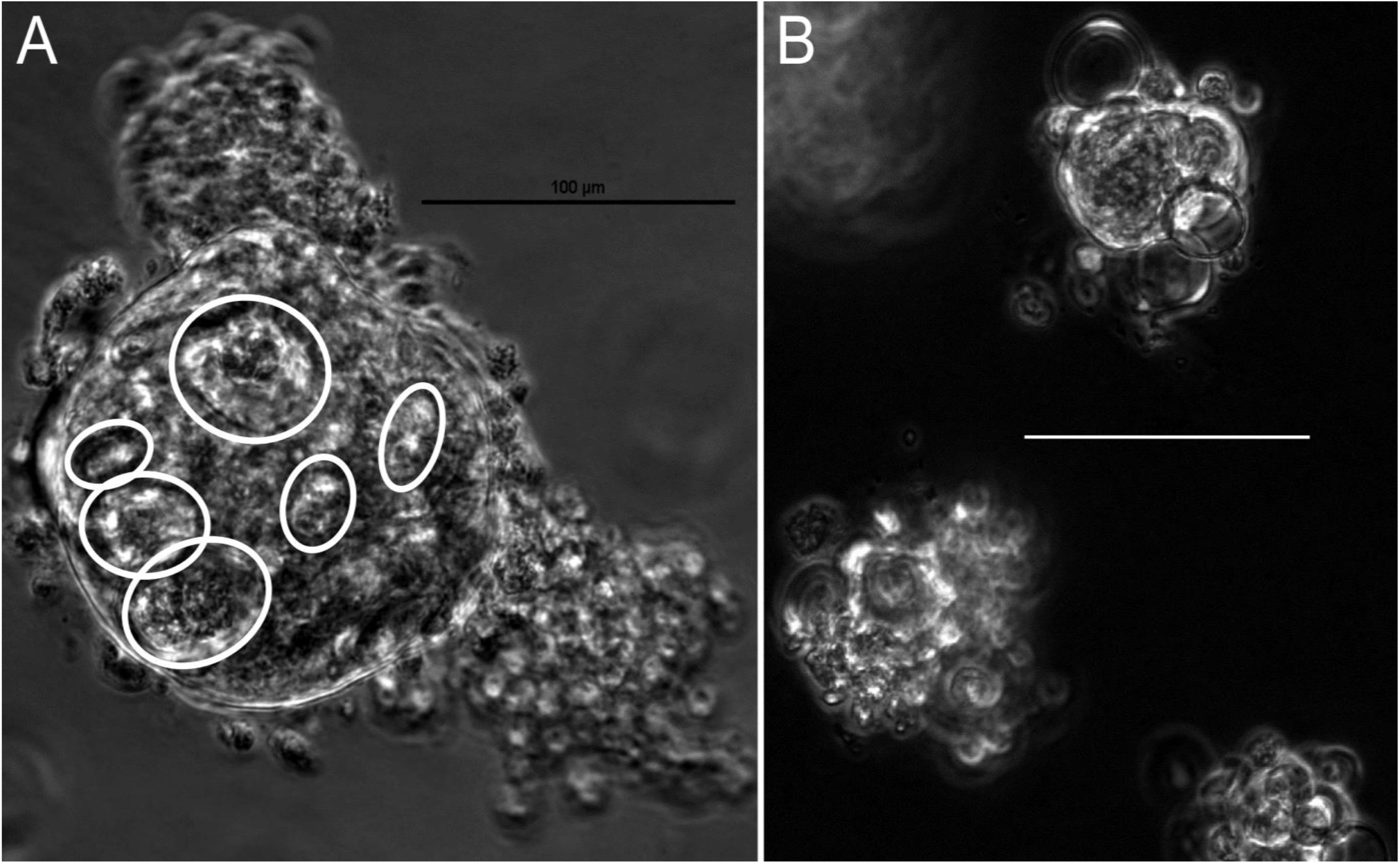
Sternum metastasis RC organoids receding in diameter after treatment with docetaxel at 1 µM. (A) Crypts are clearly visible on this image and even if the organoid edges are still intact, they appear very uneven after 48 hours of docetaxel treatment. (B) After additional 13 days of drug treatment, i.e., 15 days in total, many dead cancer cells are visible around the receded organoid. Scale bars equal 100 µm. Phase contrast microscopy, 20x magnification.

The state of persistence was initially identified in bacterial antibiotic resistance and it is a non-mutational mechanism of acquired drug resistance (Fisher et al., 2017) during which a small population (<5%) of the cells become quiescent. In cancer, the quiescent surviving persister cells minimize the rates of proliferation and, thus, become not susceptible to treatment with mitotic inhibitors and targeted therapies. This reversible process of becoming drug-tolerant is linked to upregulation of mesenchymal markers and downregulation of epithelial markers in the persister cells. During drug holiday, the cells revert to normal metabolism and expand their population, while becoming again susceptible to drug treatment. This cycle repeats multiple times during patient treatment. While the tumor is under drug pressure, however, the few drug-tolerant persister cells pay a metabolic price and become temporarily vulnerable to GPX4 inhibition (Hangauer et al., 2017). Targeting these persister cells eliminates residual disease and, thus, impedes tumor relapse. Going forward, we envisage to use persister cells derived from tumor organoids as a discovery platform to identify therapeutic agents that can eliminate residual persisters that remain after initial successful cytotoxic drug treatment.

#### Analysis of the Texture of Drug-Treated Cancer Cells

During the first few days of drug treatment, even shedding dead cells, the rectal organoids retained their distinct morphology (Fig. 7, Fig. 8). One possible approach to evaluate the variety of changes in shape and surface patterns of whole (treatment-resistant) organoids, after drug treatment, which can also be applied to the analysis of changes in MM histone morphology after acetylation, can be accomplished by morphology and texture analysis of organoid images. Such quantitative readout can be used as a biomarker for clinical response therapy in grayscale value images of patient-derived tumor organoids (Fig. 7, Fig. 8).

Patient organoids are considered sensitive to treatment when multiple image features’ changes are statistically significant, such as the shift in metrics related to the organoid edge and the contraction or oblongation of the organoid shape. Using this method, we are correlating changes in organoid images with clinical response to therapy, in order to develop a model predictive of drug response. To develop this computational platform, we started by analyzing MT cytoskeleton changes in a panel of docetaxel-sensitive (MKN-45, SNU-1, TMK-1, AZ-521) and resistant (MKN-7, SCH, Hs746T) cell lines. In this context, we examined 21 types of changes, termed features, between cells at baseline and after docetaxel treatment (Matov, 2024f).

A key to the success of an automated system for organoid pattern analysis is the design of suitable numerical descriptors (features) that capture essential information from images without being overly sensitive to variations induced by organoid shape or orientation. Many types of features have been investigated in the field of computer vision and we selected specific features to describe and quantify the changes in organoids during drug treatment, such as Haralick features (angular second moment, pixel variance, pixel entropy, etc.) (Haralick, 1983) and computation of Zernike moments (shape similarity of organoid images to Zernike polynomials - 49 polynomials and 49 features) (Boland and Murphy, 2001). Additional features that can be computed pertain to organoid morphological features, organoid edge features, the morphological skeleton, and convex hull. The morphological features comprise of the number of bright objects in the organoid image, Euler number of the organoid image, average bright object size, variance of object size, to name a few. The edge related features are the fraction of above an intensity threshold pixels along the organoid edge, the homogeneity of the gradient of organoid edge intensity, homogeneity in the direction of the organoid edge, and other. The morphological skeleton features are the average length of the morphological skeleton of the organoids, the ratio of the number of branch points in the morphological skeleton to the overall length of the skeleton, and other. The convex hull’s features consist of organoid eccentricity, the roundness of the organoid convex hull, and the fraction of the organoid convex hull pixels above an intensity threshold.

Our results after docetaxel treatment show (Fig. 9) that in populations of sensitive gastric cancer (GC) cells a high number of (19%-48% of all) features undergo statistically significant shifts (Galletti et al., 2013; Galletti et al., 2014). This shows that an affected MT network correlates with drug-target engagement and response. Moreover, sensitive cells exhibit changes in different subsets of features suggesting different underlying mechanisms of docetaxel sensitivity. Oppositely, after treatment of resistant cells with docetaxel, a change in a very few (0%-14%) of the features was a hallmark of resistance (Matov, 2024f).

**Figure 9.**
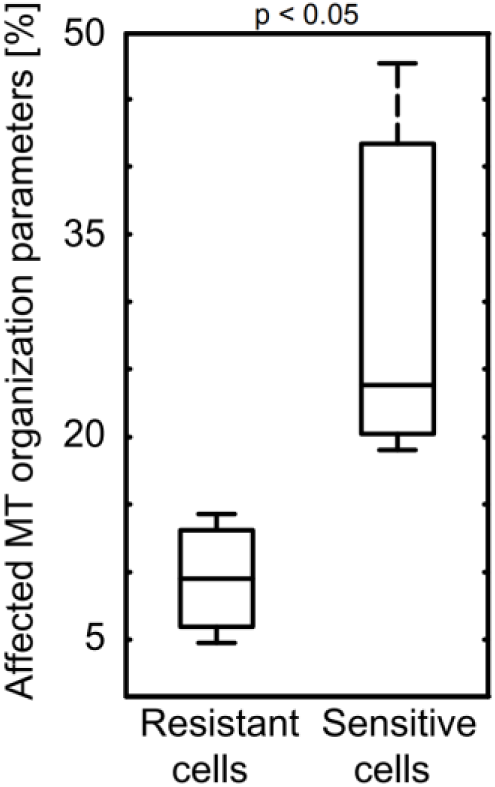
MT organization is affected in drug-sensitive GC cells. After treatment with docetaxel, up 14% of MT organization parameters were affected in resistant cell lines, while this number was up to 48% in sensitive cell lines. Images were acquired at 63x magnification.

#### Elimination of Residue Disease and Preventing Relapse

We also derived drug-tolerant persister cells in PC organoids (Gao et al., 2014), which were obtained at MSKCC from tumor resection at a retroperitoneal lymph node after receiving prior therapy with androgen deprivation therapy, bicalutamide, docetaxel, and carboplatin, i.e., the tumor was refractory to both hormonal and systemic therapy. We tested therapeutic treatments to kill these persisters (Matov, 2025). Due to a disabled antioxidant program, persister cells are unable to tolerate lipid hydroperoxide accumulation and are susceptible to ferroptotic drugs (Hangauer et al., 2017). To achieve this end, we performed experiments showing that cells derived from two to seven weeks of docetaxel treatment are sensitive to the novel compounds RSL3 and ML210, both small molecule GPX4 inhibitors.

We, therefore, sought to test if the small molecule ML210 is selectively toxic to persister cells without inducing significant toxicity in parental organoid cells. Organoids with diameter 40-80 µm were plated in 12 wells and half of the wells were treated for five weeks with ML210 at 5 µM. After five weeks of organoid culture and reaching a phase of exponential growth, the organoids had diameter size of 60-600 µm (Fig. 10). The differences in numbers of viable cells between the untreated organoids and the ML210-treated wells were not statistically significant, which demonstrated that ML210 was not toxic to the parental tumor organoid cells.

**Figure 10.**
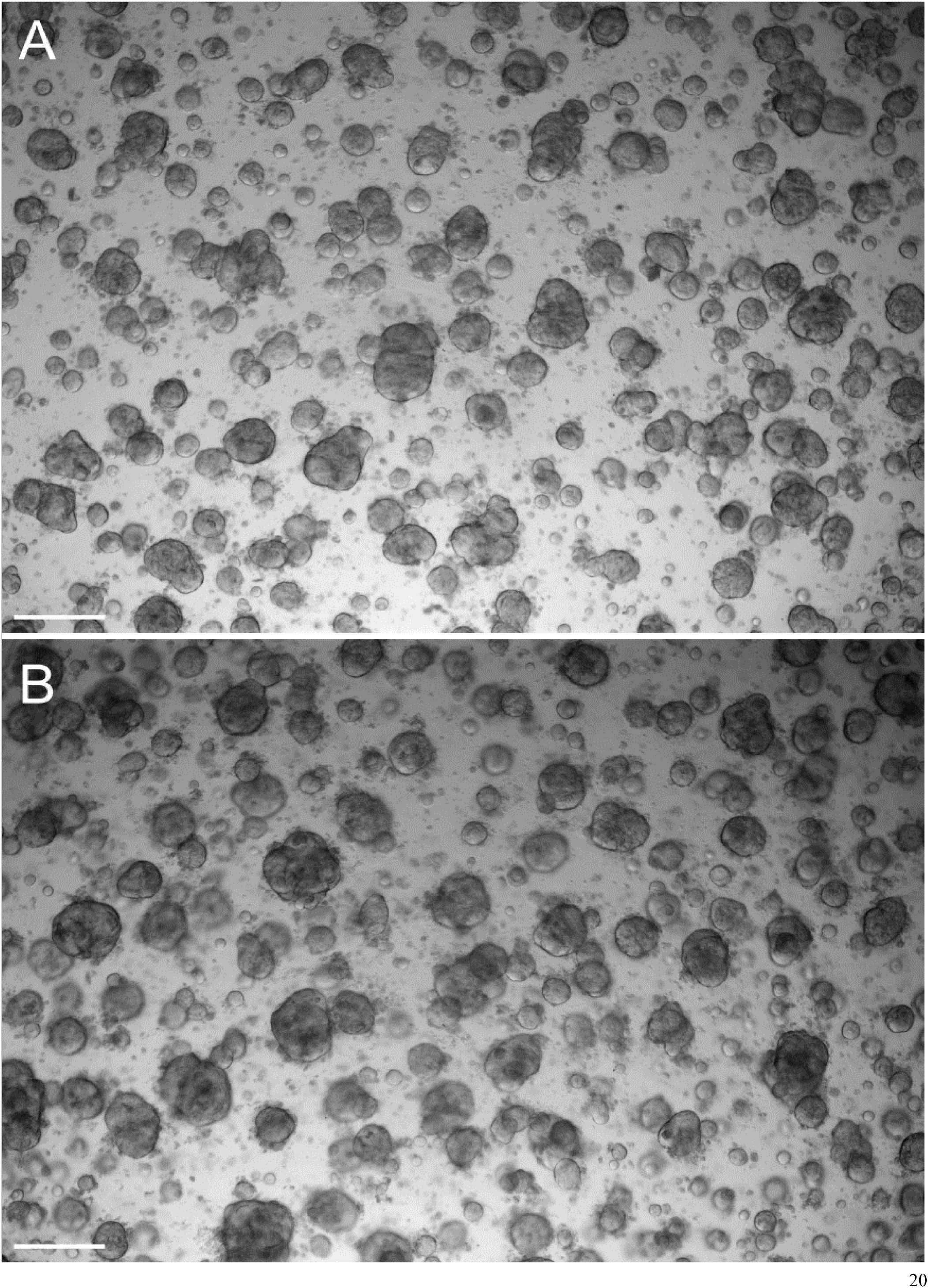
Retroperitoneal lymph node metastasis patient-derived prostate cancer organoids after no treatment and after treatment with a ferroptosis inhibitor only. Organoids with diameter 40-80 µm were plated in 12 wells and half of the wells were treated for five weeks with ML210 at 5 µM. After five weeks of culturing, the organoids had a diameter size of 60-600 µm. Overall, the differences in numbers of viable cells between the control cells and the ML210 treated cells were not statistically significant, which demonstrates that ML210 requires docetaxel-induced persister state to be toxic to the parental tumor organoid cells. (A) Image showing the organoids after five weeks with no drug added. (B) Image showing organoids after five weeks treated with ML210 only. Scale bars equal 1 mm. Transmitted light microscopy, 4x magnification.

We then plated organoids with diameter 40-80 µm in 12 wells of which four were treated with docetaxel at 100 nM for seven weeks, four wells were treated for two weeks with docetaxel at 100 nM followed by a five-week co-treatment with docetaxel at 100 nM and ML210 at 5 µM, and four wells were treated for two weeks with docetaxel at 100 nM followed by a five-week co-treatment with docetaxel at 100 nM, ML210 at 5 µM, and ferrostatin-1 at 2 µM (Fig. 11). The organoids had a diameter of 40-80 µm at baseline and the two weeks treatment with docetaxel eliminated the vast majority of the organoid cells and induced a persister state in the residual cancer cells. A follow-up co-treatment with the GPX4 inhibitor ML210 reduced the number of viable residual cells two-fold in median values (a median value of ATP luminescence of 3,000 after seven weeks of docetaxel treatment was reduced to a 1,500 median value). We measured a significant decrease of 63% in mean ATP luminescence values (p < 0.05) with only about a third of the persister cells remaining viable after ML210 co-treatment when we compared the numbers of viable cells after seven weeks of docetaxel-only treatment. Further, co-treatment with the lipophilic antioxidants ferrostatin-1 rescued the persister cells from the toxic effects of GPX4 inhibition and we measured a 44% increase in mean cell viability in the wells for which we also added ferrostatin-1 at 2 µM in comparison to the viability of the cells co-treated with ML210 only; however, the median ATP luminescence values for this experiment remained the same, which demonstrates a partial rescue (Matov, 2025).

**Figure 11.**
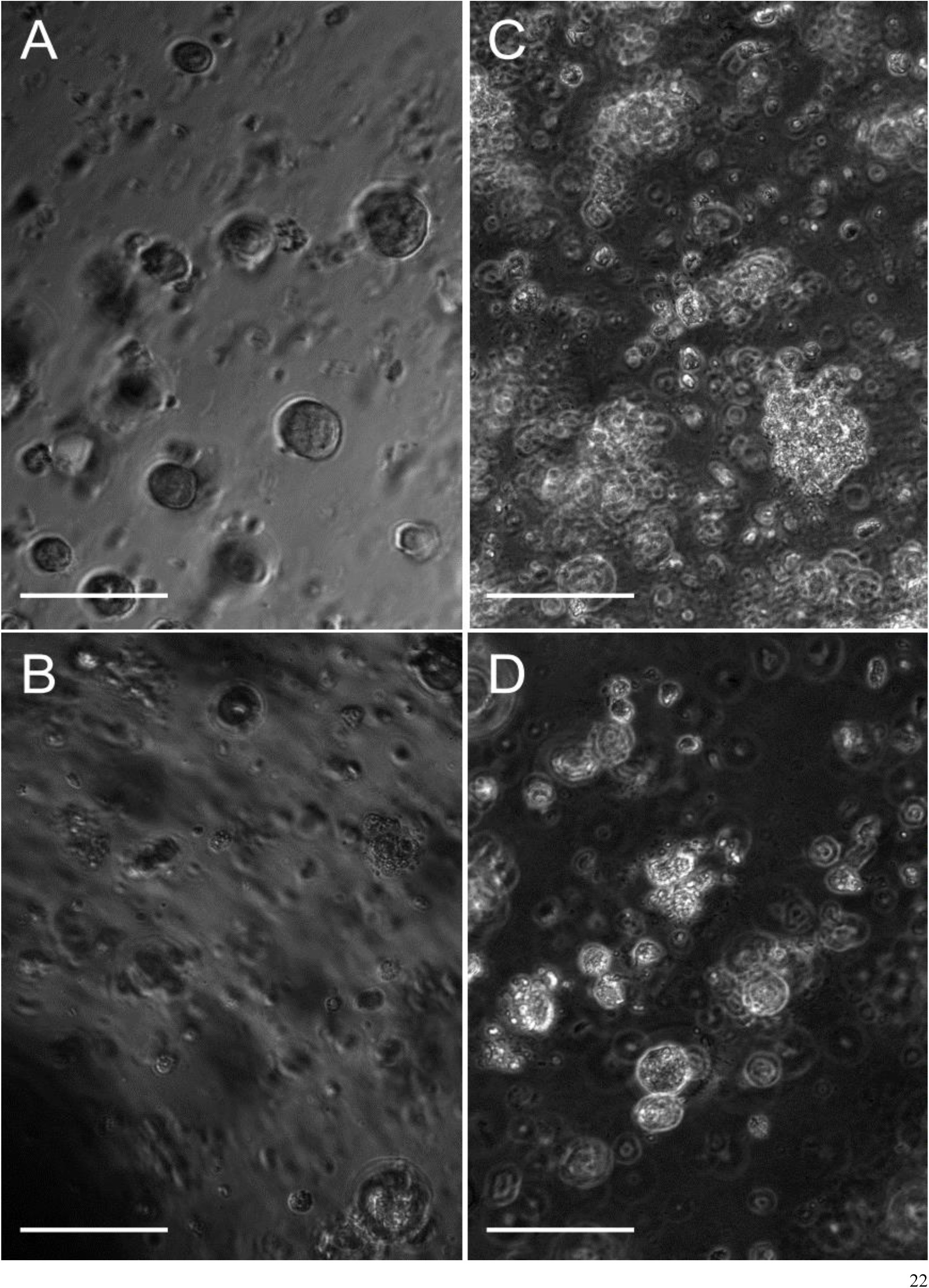
Retroperitoneal lymph node metastasis patient-derived prostate cancer organoids before and after treatment with a cytotoxic drug in combination with a ferroptosis-inducing small molecule with or without added rescue drug. (A) Image of organoids at baseline. (B) Image showing the effects on organoids of treatment with 100 nM docetaxel. (C) Image showing dead organoid cells in Matrigel after combination treatment with docetaxel at 100 nM and ML210 at 5 µM. (D) Image taken after treatment with docetaxel at 100 nM and ML210 at 5 µM together with the rescue drug ferrostatin-1 at 2 µM shows very small organoids. Scale bar equals 100 µm. Phase contrast microscopy, 20x magnification.

This result suggests that higher levels of ferrostatin-1 could rescue more efficiently the persister cells from the toxic effects of GPX4 inhibition. We also observed extensive Matrigel damage in some of the wells with ferrostatin-1, which likely contributed to the very low cell viability numbers in these particular affected wells; the Matrigel damage likely contributed also to the overall high variability and low statistical significance (p = 0.05) of the rescue experiment.

Inducing ferroptosis is a viable strategy for the elimination of residual disease in MM as well (Logie et al., 2021). The development of a real-time computer vision system (Matov, 2024f), which can reliably evaluate drug action *ex vivo* will provide an additional platform for functional testing of putative compounds and combination therapies, including those exploiting vulnerabilities in cancer cells, like the susceptibility to ferroptosis-inducing small molecules, to eliminate residual disease. Quantitative imaging could be displayed during tumor board meetings, which would allow the discussion of tentative regimens based on the supporting information provided by the microscopic evaluation of patient-derived cultures.

## DISCUSSION

Our objective is to dissect the changes in the MM cell after treatments at a microscopic level. We will use an approach for *ex vivo* anticipation of drug resistance based on the (i) *ex vivo* evaluation of IC50 values for MM cells treated in a microfluidics device and (ii) changes in live-cell behavior in terms of morphology (Matov and Modiri, 2024) and dynamics (Matov, 2024d) of cellular, molecular markers over 24 hours or longer. In this context, we will rely on a statistical analysis of large populations of molecular markers over an extended period of time to distinguish between disease phenotypes, which otherwise appear similar based on the currently utilized patient stratification methods.

### Patient-Derived MM Models

To utilize patient MM cells, the model system we propose can be based on cells extracted from BM biopsies. It can utilize different marrow cellular components to recreate the tumor microenvironment in the lab. Through flow cytometry, several cell types can be selected, based on surface antigens such as CD138 for myeloma, CD34 for stem cells, and CD105 for mesenchymal cells. A BM biopsy could yield about ten million cells and after FACS sorting, one expects to collect about 200k MM cells, 20k stem cells, <100k mesenchymal cells. Patient samples can be obtained by overnight shipping of fresh biopsies at room temperature. At the biopsy site, one would immediately isolate the BM mononuclear cells and then sort for the three markers. The BM cells are suspension cells with diameters of about seven to 15 µm (Orkin and Zon, 2008), and their exosomes have sizes between 20 and 100 nm (Thery et al., 2018). MM/BM cells are extremely sensitive to *ex vivo* culture and die (100% rate) when plated in a standard 2D culture dish. To culture them in the lab, there are culture methods that recreate their 3D microenvironment. To provide a similar environment, we will culture them on hFOB 1.19 cells, which mimic the human bone, in a specialized microfluidics device. The BM culture medium consists of RPMI with L-glutamine, patient plasma, CaCl2, sodium succinate, hydrocortisone, heparin (Zhang et al., 2015), and one should allow for 48 hours of metabolic recovery after the biopsy before an *ex vivo* drug treatment. After plating patient cells in microfluidics, about 70% of them will survive (Zhang et al., 2014). To increase the rates of patient-cell survival after plating further, we will test additional media components based on novel media formulations to improve the cell viability (Aleman et al., 2019). Lentiviral-mediated expression of the fluorescent exosome markers (Lotvall et al., 2014) CD63-RFP, CD9-GFP, and CD81-YFP can be used to mark exosomes for visualization. A combination of the three tetraspanins can be used to analyze exosomes of different origins, enriched in different, transmembrane or lipid-bound, extracellular proteins.

### Analysis of Drug Susceptability

Optimal drug selection is a challenging task and has been the topic of multiple investigations (Belloni et al., 2018; Candini et al., 2019; Geisler, 2019), and it has become clear that gene expression profile alone is inadequate in predicting therapeutic response (Amin et al., 2014). An image analysis system for drug selection in MM (Silva et al., 2017) has been shown to automatedly enumerate the dead cells using brightfield microscopy at a low spatial and temporal resolution, but *ex vivo* systems for high spatial and temporal resolution analysis are still missing. Answers about the potential efficacy of a drug can, thus, be obtained by using software systems that measure, in great detail, the effects of a regimen on its cellular targets (Matov, 2024d). Molecular manipulations of living patient-derived cells *ex vivo* can reveal which secondary mechanisms would be activated after a particular drug intervention. To achieve this end, the tracks obtained during computational motion analysis can be used for disease classification and patient stratification purposes. Depending on the methods used, the resulting analysis images may consist of either flow vectors (Matov, 2024g; Matov and Bacconi, 2024) or full trajectories (Applegate et al., 2011; Matov et al., 2010) and can be classified into sensitive or resistant using artificial intelligence (AI) methods. Tracking data image stacks, in the thousands, with tracks of the exosomes visualized, can be utilized for disease classification by training a generative transformer (Ren et al., 2024), or another large language model, which takes image patterns as input (Horiuchi et al., 2024). Such computational clinical research can aid the discovery of an optimal drug regimen selection by assigning a sensitive or resistant label to every patient cell analyzed.

### Real-Time Motion Tracking

Our approach for the evaluation of cytoskeletal modulation consisted of analysis of labeled MT tips (Matov, 2024b; Matov, 2024c; Matov, 2024d) after image acquisition and once the image dataset is transferred from the microscope to a computational machine (Vid. 2 shows an example with a colon cancer (CC) organoid we imaged and analyzed at MBL, Woods Hole). To accelerate this process and improve our ability to calibrate the drug regimen, we have developed a software suite (DataSet Tracker (Matov, 2024g)) for real-time analysis designed to run on computers, smartphones, and smart glasses hardware and suitable for resource-constrained, on-the-fly computing in microscopes without internet connectivity. We will further extend the current functionality into obtaining the full trajectories by utilizing the vectors obtained in each frame, together with the associated information on feature intensity and morphology, to generate embeddings (Vaswani et al., 2017) for a generative transformer network in which the tokens are the spatial coordinates of the features we track. This will allow training of the network to associate the most likely next feature in an image sequence, similar to the way large language models generate text. Further, we will retrain a transformer network with a new set of tokens - with lists of 2D or 3D coordinates rather than words and with trajectories (lists of linked coordinates) rather than sentences of human speech, i.e., we will retrain a large language model with motion trajectory data. Having the ability to test in quantitative detail a number of drugs and combinations *ex vivo* will be critical in the treatment of pathologies for which there is no known cure. Once the specific impaired molecular mechanisms are identified for the particular patient, treatment options which correct the aberrations can be selected in real time.

Based on clinical data for resistance, this analysis will allow for the identification of an *ex vivo* clinical resistance baseline phenotype and for the characterization of the changes in this phenotype, or lack thereof, after treatments with different drug combinations of refractory disease for which currently there is no known suitable treatment regimen. As there is currently no cure for MM, we consider modulation of the cytoskeleton as a viable new option. Refractory MM can be sensitized to therapy with the use of Rho-associated protein kinase (ROCK) inhibitors (Federico et al., 2020). ROCK regulates myosin light chain phosphorylation and as a result actin activity and cellular contractility (Matov and Bacconi, 2024). We hypothesize that tubulin inhibitors can be impactful in the treatment of refractory MM, since Rho family GTPases (Matov, 2024e) have been implicated as possible targets (Ahmed et al., 2022; Mulloy et al., 2010), and that novel targets can be identified within the MT transcriptome (examples are depicted on Fig. 1, such as CLIP-170 (Rozic et al., 2015)) or other proteins involved in the regulation of MT dynamics in cancer (Matov, 2024b; Matov, 2024d), for instance GSK3β (Augello et al., 2020).

## MATERIALS AND METHODS

### Sample Processing

Small RNA were extracted from de-identified urine samples by Norgen Biotek Corp.

### Cell Culture

Organoids with diameter 40-80 µm were plated in 30 µL Matrigel drops in six wells per condition of a 24-well plate per condition and treated for two or more weeks with docetaxel with concentration of 100 nM and 1 µM to eliminate the vast majority of the organoid cells and induce a persister state in the residual cancer cells. This treatment was followed by co-treatment with docetaxel 100 nM and small molecule ML210 5 µM or docetaxel 100 nM combined with small molecule ML210 5 µM and lipophilic antioxidants ferrostatin-1 2 µM, rescue treatment. Organoids with diameter 40-80 µm were plated in 30 µL Matrigel drops in six wells per condition of a 24-well plate per condition and also treated for two or more weeks with FOLFOX (folinic acid, fluorouracil, and oxaliplatin) with concentration of 1 µM, 10 µM, and 100 µM to eliminate the vast majority of the organoid cells and induce a persister state in the residual cancer cells.

Primary and retroperitoneal lymph node metastatic prostate tumor and sternum metastatic rectal tumor tissues were dissociated to single cells using modified protocols from the Witte lab. Organoids were seeded as single cells in three 30 µL Matrigel drops in 6-well plates. Organoid medium was prepared according to modified protocols from the Clevers lab and the Chen lab.

To obtain a single cell suspension, tissues were mechanically disrupted and digested with 5 mg/ml collagenase in advanced DMEM/F12 tissue culture medium for several hours (between 2 and 12 hours, depending on the biopsy/resection performed). If this steps yielded too much contamination with non-epithelial cells, for instance during processing of primary prostate tumors, the protocol incorporated additional washes and red blood cell lysis (Goldstein et al., 2011). Single cells were then counted using a hemocytometer to estimate the number of tumor cells in the sample, and seeded in growth factor-reduced Matrigel drops overlaid with PC medium (Gao et al., 2014). With radical prostatectomy specimens, we had good success with seeding three thousand cells per 30 μl Matrigel drop, but for metastatic samples organoid seeding could reliably be accomplished with significantly less cells, in the hundreds. To derive organoids from patient CTCs, liquid biopsy samples of 40 ml peripheral blood would be collected, processed, and plated in a Matrigel-Collagen-Fibronectin matrix to form organoids similarly to the metastatic breast cancer organoids we cultured from mouse CTCs (Matov, 2024b).

To transduce organoids, we modified protocols from the Clevers lab to adapt to the specifics of prostate organoid culture (such as the significant differences in proliferation rates in comparison to colon and rectal organoids). We found out empirically that cells in mid-size organoids (60-100 µm in diameter) infect at much better rates than trypsinized single organoid cells. These were the steps we followed to express EB1ΔC-2xEGFP in organoids: (1) Add Dispase (1 mg/ml) to each well to dissolve the Matrigel at room temperature for 1 hour. (2) Spin down (at 1,000 rpm for 4 minutes) and mix organoids with 10 µl of viral particles (enough for 1 well with three Matrigel drops of 30-40 µl with organoids containing 1-2 million cells) with Y27632 ROCK inhibitor and Polybrene (1:1,000) for 30 minutes. (3) Spin the organoids with viral particles for 1 hour at 600g. (4) Leave the organoids for a 6-hour incubation. (5) Spin down and plate in Matrigel. (6) 1 hour later, add medium. We used blasticidin (1:20,000) for only one round of medium (3 days) because an increase of the density of labeled cells in the organoids reduced our ability to image MT tips with good contrast.

To evaluate the metastatic potential of the RC organoids, we performed an overnight the Boyden Chamber cell migration transwell assay (Chen, 2005).

Exosomes in MM1S cells (a gift from the Thaxton lab) were visualized by lentivirus-mediated low level expression of CD63-RFP. To generate imaging data, we performed lentiviral infection with CD63-RFP and observed the labeling signal in the red channel 16 hours later. We did not use blasticidin selection as the infection rate was over 90% and performed a 24-hour titration with dexamethasone at concentrations 60 nM, 120 nM, and 250 nM.

Docetaxel was purchased from Sigma-Aldrich. Dexamethasone was a gift from the Wiita lab. Folinic acid, fluorouracil, oxaliplatin, RSL3, ML210, and ferrostatin-1 were obtained from the McCormick lab.

### Microscopy Imaging

We imaged CD63-RFP-expressing MM1S cells by time-lapse spinning disk confocal microscopy using a 100x oil immersion objective and acquired 3D z-stacks, which consisted of 20 2D slices acquired with a 500 ms exposure time at a 2 µm vertical spacing. The lateral resolution of a 1.49 NA objective is 220 nM. The temporal frequency with which we acquired 3D image stacks was 11 seconds.

Organoids were imaged using transmitted light microscopy at 4x magnification and phase contrast microscopy at 20x magnification on a Nikon Eclipse Ti system with camera Photometrics CoolSnap HQ2.

We imaged MT tips in EB1ΔC-2xEGFP-expressing organoids (Koo et al., 2013) by time-lapse spinning disk confocal microscopy using a long working distance 60x magnification water immersion 1.45 NA objective. We acquired images at half a second intervals for a minute to collect datasets without photobleaching (Gierke and Wittmann, 2012).

### Image Analysis

All image data analysis programs and graphical representation of exosomes were developed in Matlab and R, and available upon request. The 3D SIFT Flow method for 3D displacement fields analysis computation is described and validated in (Liu et al., 2011). Image analysis programs for EB1 comet detection, MT dynamics, and graphical representation of the MT tips analysis were developed in Matlab and C/C++; the ClusterTrack method is described and validated in (Matov et al., 2010). The DataSet Tracker computer code is available for download at: https://www.github.com/amatov/ClusterTrackTubuline. The software suite for real-time image analysis was developed in the cross-platform game engine Unity in C# (Matov, 2024g). The computer code is available for download at: https://www.github.com/amatov/DataSet Tracker. OpenCV for Unity requires a license from Enox Software. The image analysis programs for PSMA, α-tubulin, DAPI, and CD45 segmentation as well as 3D AR and DAPI segmentation, and graphical representation of the results were developed in Matlab and C/C++. The wavelet transform method used, spotDetector, was described and validated in (Olivo-Marin, 2002) and the unimodal pixel intensity thresholding in (Rosin, 2001). We identified the CTC areas as connected-component labeling pixel lists in the epithelial tumor imaging channel (such as PSMA or occasionally cytokeratin) for which in the nuclear imaging channel there is a DAPI stain with a statistically representative size and a circular shape. At the same time, we required that these cells are negative in the CD45, i.e., the absence of a leukocyte marker. Similarly, to detect neutrophils and lymphocytes, we identified clusters of bright pixels in the CD45 channel for which a nuclear area is detected in the DAPI channel and the epithelial stain is not present. On multiple occasions, we detected double-positive (PSMA+/DAPI+/CD45+ or CK+/DAPI+/CD45+) and double-negative (PSMA-/DAPI+/CD45- or CK-/DAPI+/CD45-) cells, which we classified in separate bins. Drug-induced MT bundles were evaluated in terms of thickness and texture parameters in 63x images. The detection of tyrosinated tubulin in 4x images was considered to be a marker of drug sensitivity. We analyzed the cellular localization of AR in 3D image stacks, in which we performed segmentation in 35 individual 2D images and reconstructed the volume, and classified the CTCs as sensitive to drug treatment if the AR is within the volume of the nucleus and resistant if the AR is mainly in the cytoplasm. The computer code is available for download at: https://github.com/amatov/SegmentationBiomarkerCTC, https://github.com/amatov/ResistanceBiomarkerAnalysis, https://github.com/amatov/AntibodyTextureMorphology.

### Statistical Analysis

No statistical methods were used to predetermine sample size. For organoid experiments, the samples were not randomized. Statistical tests were performed in Matlab.

The ATP luminescence datasets were not sufficiently large to directly apply a Student’s *t*-test. Instead, we used a non-parametric permutation *t*-test with 400 repetitions (Hesterberg et al., 2005), which compares the bootstrapped distributions of a condition 1 versus condition 2. This test does not make any assumption about the characteristic of the tested distribution and, thus, is appropriate for application to distributions that are far from being normally distributed. In brief, for each condition 400 values are bootstrap-sampled from the data of different experimental conditions. In agreement with the central limit theorem (Laplace, 1810), the two bootstrapped distributions always follow a Gaussian distribution and, thus, could be analyzed for differences using a regular Student’s *t*-test.

## Supporting information

Transwell assay demonstrates the metastatic potential of patient RC cells.

CC organoid with labeled, detected, and tracked MT tips.

## Data Availability

All data produced in the present study are available upon reasonable request to the authors

## List of Abbreviations

AR: androgen receptor
BM: bone marrow
BMSC: bone marrow stromal cells
CLIP-170: cytoplasmic linker protein 170
CC: colon cancer
CTC: circulating tumor cell
EB1/3: microtubule end binding protein, 1/3
EB1ΔC: EB1 construct truncated at amino acid 248 that does not interact with other proteins at the microtubule end
EGFP: enhanced green fluorescent protein (2xEGFP indicates two molecules)
GC: gastric cancer
GPX4: glutathione peroxidase 4
GSK3α/β: glycogen synthase kinase 3, alpha/beta isoforms
MT: microtubule
MGUS: monoclonal gammopathy of undetermined significance
MM: multiple myeloma
PC: prostate cancer
PSMA: prostate-specific membrane antigen
ROCK: Rho-associated coiled-coil kinase
RC: rectal cancer
SMM: smoldering multiple myeloma

## Ethics Declaration

IRB (IRCM-2019-201, IRB DS-NA-001) of the Institute of Regenerative and Cellular Medicine. Ethical approval was given.

Approval of tissue requests #14-04 and #16-05 to the UCSF Cancer Center Tissue Core and the Genitourinary Oncology Program was given.

## Data Availability Statement

The datasets used and/or analyzed during the current study are available from the corresponding author upon reasonable request.

## ACKNOWLEDGEMENTS

I thank Michael Plebanek for MM1S cells and CD63-RFP virus particles, Arun Wiita and his lab for their help, Anna Celli for 3D imaging help, and JR&D Services for funding in the context of NGS data. I am grateful to the Genitourinary Tissue Utilization committee and the Genitourinary and Prostate SPORE Tissue Cores at the UCSF Cancer Center for the approval of my tissue requests #14-04 and #16-5, the Stand Up To Cancer / Prostate Cancer Foundation (SU2C/PCF) West Coast Dream Team (WCDT), the Institute of Regenerative and Cellular Medicine for issuing the Institutional Review Board protocol approval IRCM-2019-201, IRB DS-NA-001 for the observational study “Longitudinal analysis of next-generation sequencing of nucleic acids for early detection of degenerative diseases such as cardiovascular, neoplastic and diseases related to the nervous system”, and James Faber for his feedback regarding the protocol and the process of approval. The patient blood samples analyzed were from clinical studies with IRB protocols 0804009740 and 0707009283 at Cornell Medicine. A preprint of this paper is available at medRxiv (Matov, 2024a).

## SUPPLEMENTARY MATERIALS

Video 1 – Transwell assay demonstrates the metastatic potential of patient RC cells. The experiment was conducted overnight and images were acquired every 10 minutes. https://vimeo.com/1057363291/27aa30bedd

Video 2 – CC organoid with labeled, detected, and tracked MT tips. Only persistent trajectories of at least four frames are shown and shorter ones were omitted as potential noise artifacts. The overlaid trajectories (in red) are displayed only for the duration of MT polymerization. https://vimeo.com/1035848286/6f3a2c0f0e

## REFERENCES

Ahmed, S., S. Mazumder, L. Wiesen, N. Sharma, U.K. Mukherjee, S.K. Kumar, L.B. Baughn, B.G. Van Ness, and A.K. Mitra. 2022. Validation of a Novel Rac1 Inhibitor As Potent Secdrug Against Activating Ras Mutant and Drug-Resistant Multiple Myeloma. Blood. 140:4249–4250.

Aleman, J., S.K. George, S. Herberg, M. Devarasetty, C.D. Porada, A. Skardal, and G. Almeida-Porada. 2019. Deconstructed Microfluidic Bone Marrow On-A-Chip to Study Normal and Malignant Hemopoietic Cell-Niche Interactions. Small:e1902971.

Amin, S.B., W.K. Yip, S. Minvielle, A. Broyl, Y. Li, B. Hanlon, D. Swanson, P.K. Shah, P. Moreau, B. van der Holt, M. van Duin, F. Magrangeas, P. Pieter Sonneveld, K.C. Anderson, C. Li, H. Avet-Loiseau, and N.C. Munshi. 2014. Gene expression profile alone is inadequate in predicting complete response in multiple myeloma. Leukemia. 28:2229–2234.

Anderson, K.C. 2009. Proteasome inhibitors in multiple myeloma. Semin Oncol. 36:S20–26.

Applegate, K.T., S. Besson, A. Matov, M.H. Bagonis, K. Jaqaman, and G. Danuser. 2011. plusTipTracker: Quantitative image analysis software for the measurement of microtubule dynamics. J Struct Biol. 176:168–184.

Attal, M., V. Lauwers-Cances, C. Hulin, X. Leleu, D. Caillot, M. Escoffre, B. Arnulf, M. Macro, K. Belhadj, L. Garderet, M. Roussel, C. Payen, C. Mathiot, J.P. Fermand, N. Meuleman, S. Rollet, M.E. Maglio, A.A. Zeytoonjian, E.A. Weller, N. Munshi, K.C. Anderson, P.G. Richardson, T. Facon, H. Avet-Loiseau, J.L. Harousseau, and P. Moreau. 2017. Lenalidomide, Bortezomib, and Dexamethasone with Transplantation for Myeloma. The New England journal of medicine. 376:1311–1320.

Augello, G., M.R. Emma, A. Cusimano, A. Azzolina, G. Montalto, J.A. McCubrey, and M. Cervello. 2020. The Role of GSK-3 in Cancer Immunotherapy: GSK-3 Inhibitors as a New Frontier in Cancer Treatment. Cells. 9.

Belloni, D., S. Heltai, M. Ponzoni, A. Villa, B. Vergani, L. Pecciarini, M. Marcatti, S. Girlanda, G. Tonon, F. Ciceri, F. Caligaris-Cappio, M. Ferrarini, and E. Ferrero. 2018. Modeling multiple myeloma-bone marrow interactions and response to drugs in a 3D surrogate microenvironment. Haematologica. 103:707–716.

Boland, M.V., and R.F. Murphy. 2001. A neural network classifier capable of recognizing the patterns of all major subcellular structures in fluorescence microscope images of HeLa cells Bioinformatics. 17:1213–1223.

Candini, O., G. Grisendi, E.M. Foppiani, M. Brogli, B. Aramini, V. Masciale, C. Spano, T. Petrachi, E. Veronesi, P. Conte, G. Mari, and M. Dominici. 2019. A Novel 3D In Vitro Platform for Pre-Clinical Investigations in Drug Testing, Gene Therapy, and Immuno-oncology. Sci Rep. 9:7154.

Chandra, R., K. Jain, R.V. Deo, and S. Cripps. 2019. Langevin-gradient parallel tempering for Bayesian neural learning. Neurocomputing. 359:315–326.

Chari, A., D.T. Vogl, M. Gavriatopoulou, A.K. Nooka, A.J. Yee, C.A. Huff, P. Moreau, D. Dingli, C. Cole, S. Lonial, M. Dimopoulos, A.K. Stewart, J. Richter, R. Vij, S. Tuchman, M.S. Raab, K.C. Weisel, M. Delforge, R.F. Cornell, D. Kaminetzky, J.E. Hoffman, L.J. Costa, T.L. Parker, M. Levy, M. Schreder, N. Meuleman, L. Frenzel, M. Mohty, S. Choquet, G. Schiller, R.L. Comenzo, M. Engelhardt, T. Illmer, P. Vlummens, C. Doyen, T. Facon, L. Karlin, A. Perrot, K. Podar, M.G. Kauffman, S. Shacham, L. Li, S. Tang, C. Picklesimer, J.R. Saint-Martin, M. Crochiere, H. Chang, S. Parekh, Y. Landesman, J. Shah, P.G. Richardson, and S. Jagannath. 2019. Oral Selinexor-Dexamethasone for Triple-Class Refractory Multiple Myeloma. The New England journal of medicine. 381:727–738.

Chen, B.C., W.R. Legant, K. Wang, L. Shao, D.E. Milkie, M.W. Davidson, C. Janetopoulos, X.S. Wu, J.A. Hammer, 3rd, Z. Liu, B.P. English, Y. Mimori-Kiyosue, D.P. Romero, A.T. Ritter, J. Lippincott-Schwartz, L. Fritz-Laylin, R.D. Mullins, D.M. Mitchell, J.N. Bembenek, A.C. Reymann, R. Bohme, S.W. Grill, J.T. Wang, G. Seydoux, U.S. Tulu, D.P. Kiehart, and E. Betzig. 2014. Lattice light-sheet microscopy: imaging molecules to embryos at high spatiotemporal resolution. Science (New York, N.Y.). 346:1257998.

Chen, H.-C. 2005. Boyden Chamber Assay. *In* Cell Migration: Developmental Methods and Protocols. J.-L. Guan, editor. Humana Press, Totowa, NJ. 15–22.

Chen, X., R. Kang, G. Kroemer, and D. Tang. 2021. Broadening horizons: the role of ferroptosis in cancer. Nat Rev Clin Oncol. 18:280–296.

Child, J.A., G.J. Morgan, F.E. Davies, R.G. Owen, S.E. Bell, K. Hawkins, J. Brown, M.T. Drayson, and P.J. Selby. 2003. High-dose chemotherapy with hematopoietic stem-cell rescue for multiple myeloma. The New England journal of medicine. 348:1875–1883.

Choudhry, P., D. Galligan, and A.P. Wiita. 2018. Seeking Convergence and Cure with New Myeloma Therapies. Trends in cancer. 4:567–582.

Cohen, Y.C., H. Magen, M. Gatt, M. Sebag, K. Kim, C.K. Min, E.M. Ocio, S.S. Yoon, M.P. Chu, P. Rodríguez-Otero, I. Avivi, N.A. Quijano Cardé, A. Kumar, M. Krevvata, M.R. Peterson, L. Di Scala, E. Scott, B. Hilder, J. Vanak, A. Banerjee, A. Oriol, D. Morillo, and M.V. Mateos. 2025. Talquetamab plus Teclistamab in Relapsed or Refractory Multiple Myeloma. The New England journal of medicine. 392:138–149.

Dimopoulos, M.A., P.M. Voorhees, F. Schjesvold, Y.C. Cohen, V. Hungria, I. Sandhu, J. Lindsay, R.I. Baker, K. Suzuki, H. Kosugi, M.D. Levin, M. Beksac, K. Stockerl-Goldstein, A. Oriol, G. Mikala, G. Garate, K. Theunissen, I. Spicka, A.K. Mylin, S. Bringhen, K. Uttervall, B. Pula, E. Medvedova, A.J. Cowan, P. Moreau, M.V. Mateos, H. Goldschmidt, T. Ahmadi, L. Sha, A. Cortoos, E.G. Katz, E. Rousseau, L. Li, R.M. Dennis, R. Carson, and S.V. Rajkumar. 2024. Daratumumab or Active Monitoring for High-Risk Smoldering Multiple Myeloma. The New England journal of medicine.

Facon, T., M.A. Dimopoulos, X.P. Leleu, M. Beksac, L. Pour, R. Hájek, Z. Liu, J. Minarik, P. Moreau, J. Romejko-Jarosinska, I. Spicka, V.I. Vorobyev, B. Besemer, T. Ishida, W. Janowski, S. Kalayoglu-Besisik, G. Parmar, P. Robak, E. Zamagni, H. Goldschmidt, T.G. Martin, S. Manier, M. Mohty, C. Oprea, M.F. Brégeault, S. Macé, C. Berthou, D. Bregman, Z. Klippel, and R.Z. Orlowski. 2024. Isatuximab, Bortezomib, Lenalidomide, and Dexamethasone for Multiple Myeloma. The New England journal of medicine. 391:1597–1609.

Facon, T., S. Kumar, T. Plesner, R.Z. Orlowski, P. Moreau, N. Bahlis, S. Basu, H. Nahi, C. Hulin, H. Quach, H. Goldschmidt, M. O’Dwyer, A. Perrot, C.P. Venner, K. Weisel, J.R. Mace, N. Raje, M. Attal, M. Tiab, M. Macro, L. Frenzel, X. Leleu, T. Ahmadi, C. Chiu, J. Wang, R. Van Rampelbergh, C.M. Uhlar, R. Kobos, M. Qi, and S.Z. Usmani. 2019. Daratumumab plus Lenalidomide and Dexamethasone for Untreated Myeloma. The New England journal of medicine. 380:2104–2115.

Farhy, C., S. Hariharan, J. Ylanko, L. Orozco, F.Y. Zeng, I. Pass, F. Ugarte, E.C. Forsberg, C.T. Huang, D.W. Andrews, and A.V. Terskikh. 2019. Improving drug discovery using image-based multiparametric analysis of the epigenetic landscape. eLife. 8.

Federico, C., K. Alhallak, J. Sun, K. Duncan, F. Azab, G.P. Sudlow, P. de la Puente, B. Muz, V. Kapoor, L. Zhang, F. Yuan, M. Markovic, J. Kotsybar, K. Wasden, N. Guenthner, S. Gurley, J. King, D. Kohnen, N.N. Salama, D. Thotala, D.E. Hallahan, R. Vij, J.F. DiPersio, S. Achilefu, and A.K. Azab. 2020. Tumor microenvironment-targeted nanoparticles loaded with bortezomib and ROCK inhibitor improve efficacy in multiple myeloma. Nature communications. 11:6037.

Fisher, R.A., B. Gollan, and S. Helaine. 2017. Persistent bacterial infections and persister cells. Nature reviews. Microbiology. 15:453–464.

Galletti, G., K. Cleveland, A. Matov, J.E. Hayes, R.J. Klein, D.C. Hassane, L.V. Nicacio, and P. Giannakakou. 2013. Clinical and preclinical evaluation of taxane sensitivity in gastric cancer (GC): Relevance of GC histology. Journal of Clinical Oncology. 31:37–37.

Galletti, G., K. Cleveland, C. Zhang, A. Gjyrezi, A. Matov, D. Betel, M.A. Shah, and P. Giannakakou. 2014. Elucidating the molecular basis of intrinsic taxane resistance in gastric cancer. Cancer Research. 74:897.

Gao, D., I. Vela, A. Sboner, P.J. Iaquinta, W.R. Karthaus, A. Gopalan, C. Dowling, J.N. Wanjala, E.A. Undvall, V.K. Arora, J. Wongvipat, M. Kossai, S. Ramazanoglu, L.P. Barboza, W. Di, Z. Cao, Q.F. Zhang, I. Sirota, L. Ran, T.Y. MacDonald, H. Beltran, J.M. Mosquera, K.A. Touijer, P.T. Scardino, V.P. Laudone, K.R. Curtis, D.E. Rathkopf, M.J. Morris, D.C. Danila, S.F. Slovin, S.B. Solomon, J.A. Eastham, P. Chi, B. Carver, M.A. Rubin, H.I. Scher, H. Clevers, C.L. Sawyers, and Y. Chen. 2014. Organoid cultures derived from patients with advanced prostate cancer. Cell. 159:176–187.

Garfall, A.L., M.V. Maus, W.T. Hwang, S.F. Lacey, Y.D. Mahnke, J.J. Melenhorst, Z. Zheng, D.T. Vogl, A.D. Cohen, B.M. Weiss, K. Dengel, N.D. Kerr, A. Bagg, B.L. Levine, C.H. June, and E.A. Stadtmauer. 2015. Chimeric Antigen Receptor T Cells against CD19 for Multiple Myeloma. The New England journal of medicine. 373:1040–1047.

Gebert, L.F., M.A. Rebhan, S.E. Crivelli, R. Denzler, M. Stoffel, and J. Hall. 2014. Miravirsen (SPC3649) can inhibit the biogenesis of miR-122. Nucleic acids research. 42:609–621.

Geisler, J.G. 2019. 2,4 Dinitrophenol as Medicine. Cells. 8.

Gierke, S., and T. Wittmann. 2012. EB1-recruited microtubule +TIP complexes coordinate protrusion dynamics during 3D epithelial remodeling. Current biology : CB. 22:753–762.

Goldstein, A.S., J.M. Drake, D.L. Burnes, D.S. Finley, H. Zhang, R.E. Reiter, J. Huang, and O.N. Witte. 2011. Purification and direct transformation of epithelial progenitor cells from primary human prostate. Nat Protoc. 6:656–667.

Greenstein, S., N.L. Krett, Y. Kurosawa, C. Ma, D. Chauhan, T. Hideshima, K.C. Anderson, and S.T. Rosen. 2003. Characterization of the MM.1 human multiple myeloma (MM) cell lines: a model system to elucidate the characteristics, behavior, and signaling of steroid-sensitive and -resistant MM cells. Exp Hematol. 31:271–282.

Hangauer, M.J., V.S. Viswanathan, M.J. Ryan, D. Bole, J.K. Eaton, A. Matov, J. Galeas, H.D. Dhruv, M.E. Berens, S.L. Schreiber, F. McCormick, and M.T. McManus. 2017. Drug-tolerant persister cancer cells are vulnerable to GPX4 inhibition. Nature. 551:247–250.

Haralick, R.M. 1983. Ridges and Valleys on Digital Images. Computer Vision Graphics and Image Processing. 22:28–38.

Hesterberg, T.C., D.S. Moore, S. Monaghan, A. Clipson, and R. Epstein. 2005. Bootstrap Methods and Permutation Tests. In Introduction to the Practice of Statistics. W.H. Freeman and Co.

Hideshima, T., J. Qi, R.M. Paranal, W. Tang, E. Greenberg, N. West, M.E. Colling, G. Estiu, R. Mazitschek, J.A. Perry, H. Ohguchi, F. Cottini, N. Mimura, G. Gorgun, Y.T. Tai, P.G. Richardson, R.D. Carrasco, O. Wiest, S.L. Schreiber, K.C. Anderson, and J.E. Bradner. 2016. Discovery of selective small-molecule HDAC6 inhibitor for overcoming proteasome inhibitor resistance in multiple myeloma. Proceedings of the National Academy of Sciences of the United States of America. 113:13162–13167.

Horiuchi, D., H. Tatekawa, T. Oura, S. Oue, S.L. Walston, H. Takita, S. Matsushita, Y. Mitsuyama, T. Shimono, Y. Miki, and D. Ueda. 2024. Comparing the Diagnostic Performance of GPT-4-based ChatGPT, GPT-4V-based ChatGPT, and Radiologists in Challenging Neuroradiology Cases. Clinical neuroradiology.

Hou, R.Z., C. Chen, R. Sukthankar, and M. Shah. 2019. An Efficient 3D CNN for Action/Object Segmentation in Video. British Machine Vision Conference.

Huang, F.I., Y.W. Wu, T.Y. Sung, J.P. Liou, M.H. Lin, S.L. Pan, and C.R. Yang. 2019. MPT0G413, A Novel HDAC6-Selective Inhibitor, and Bortezomib Synergistically Exert Anti-tumor Activity in Multiple Myeloma Cells. Frontiers in oncology. 9:249.

Koo, B.K., V. Sasselli, and H. Clevers. 2013. Retroviral gene expression control in primary organoid cultures. Current protocols in stem cell biology. 27:Unit 5A.6.

Landgren, O., P. Sonneveld, A. Jakubowiak, M. Mohty, K.S. Iskander, K. Mezzi, and D.S. Siegel. 2019. Carfilzomib with immunomodulatory drugs for the treatment of newly diagnosed multiple myeloma. Leukemia.

Laplace, P.-S. 1810. Memoire sur les approximations des formules qui sont fonctions de tres grands nombres et sur leur application aux probabilites. Mémoires de l’Académie royale des sciences:353–415.

Laubach, J. 2019. Initial Therapy in Older Patients with Multiple Myeloma. The New England journal of medicine. 380:2172–2173.

Lauffart, B., S.J. Howell, J.E. Tasch, J.K. Cowell, and I.H. Still. 2002. Interaction of the transforming acidic coiled-coil 1 (TACC1) protein with ch-TOG and GAS41/NuBI1 suggests multiple TACC1-containing protein complexes in human cells. Biochem J. 363:195–200.

Lieberkühn, J.N. 1782. De Valvula Coli et Usu Processus Vermicularis. Dissertationes Quatuor.

Lindeberg, T. 1993. Detecting Salient Blob-Like Image Structures and Their Scales with a Scale-Space Primal Sketch: A Method for Focus-of-Attention. International Journal of Computer Vision 11.

Lindeberg, T. 1998. Feature detection with automatic scale selection. International Journal of Computer Vision. 30:79–116.

Liu, C., J. Yuen, A. Torralba, and W.T. Freeman. 2011. SIFT Flow: Dense Correspondence across Scenes and Its Applications IEEE Transactions on Pattern Analysis and Machine Intelligence 33:978–994.

Logie, E., B. Van Puyvelde, B. Cuypers, A. Schepers, H. Berghmans, J. Verdonck, K. Laukens, L. Godderis, M. Dhaenens, D. Deforce, and W. Vanden Berghe. 2021. Ferroptosis Induction in Multiple Myeloma Cells Triggers DNA Methylation and Histone Modification Changes Associated with Cellular Senescence. International journal of molecular sciences. 22.

Lonial, S. 2010. Relapsed multiple myeloma. Hematology Am Soc Hematol Educ Program. 2010:303–309.

Lotvall, J., A.F. Hill, F. Hochberg, E.I. Buzas, D. Di Vizio, C. Gardiner, Y.S. Gho, I.V. Kurochkin, S. Mathivanan, P. Quesenberry, S. Sahoo, H. Tahara, M.H. Wauben, K.W. Witwer, and C. Thery. 2014. Minimal experimental requirements for definition of extracellular vesicles and their functions: a position statement from the International Society for Extracellular Vesicles. Journal of extracellular vesicles. 3:26913.

Lowe, D.G. 1999a. Object recognition from local scale-invariant features. IEEE International Conference on Computer Vision. 2:1150–1157.

Lowe, D.G. 1999b. Object Recognition from Local Scale-Invariant Features. International Conference on Computer Vision and Image Understanding.

Matov, A. 2024a. Analysis of Multiple Myeloma Drug Efficacy. medRxiv:2024.24311450.

Matov, A. 2024b. Computational Analysis of Treatment Resistant Cancer Cells. medRxiv:2024.24312813.

Matov, A. 2024c. Microtubule Regulation in Cancer Cells SSRN:2024.5072927.

Matov, A. 2024d. Mitosis, Cytoskeleton Regulation, and Drug Resistance in Receptor Triple Negative Breast Cancer. arXiv:2407.19112.

Matov, A. 2024e. Modulation of the Cytoskeleton for Cancer Therapy *SSRN*:2024.5061950.

Matov, A. 2024f. Quantitative Video Microscopy in Medicine. SSRN:2024.4909311.

Matov, A. 2024g. A Real-Time Suite of Biological Cell Image Analysis Software for Computers, Smartphones, and Smart Glasses, Suitable for Resource-Constrained Computing. arXiv:2407.15735.

Matov, A. 2024h. Urinary Biomarkers for Lung Cancer Detection. medRxiv:2024.24311186.

Matov, A. 2025. Microtubule Dysregulation in Prostate Cancer Metastasis to the Vertebrae SSRN:2025.5112427.

Matov, A., K. Applegate, P. Kumar, C. Thoma, W. Krek, G. Danuser, and T. Wittmann. 2010. Analysis of microtubule dynamic instability using a plus-end growth marker. Nature Methods. 7:761–768.

Matov, A., and A. Bacconi. 2024. Instantaneous Flow Analysis of Contractile Cytoskeletal Structures Affected by the Dysregulation of Kinesin and Tropomyosin Motors. bioRxiv:2024.601275.

Matov, A., and S. Modiri. 2024. Quantitative Microscopy in Medicine. medRxiv:2024.24311304.

Matulis, S.M., V.A. Gupta, A.K. Nooka, H.V. Hollen, J.L. Kaufman, S. Lonial, and L.H. Boise. 2016. Dexamethasone treatment promotes Bcl-2 dependence in multiple myeloma resulting in sensitivity to venetoclax. Leukemia. 30:1086–1093.

McIntyre, O.R. 1979. Multiple Myeloma. The New England journal of medicine. 301:193–196.

Menu, E., and K. Vanderkerken. 2022. Exosomes in multiple myeloma: from bench to bedside. Blood. 140:2429–2442.

Mollinedo, F., J. de la Iglesia-Vicente, C. Gajate, A. Estella-Hermoso de Mendoza, J.A. Villa-Pulgarin, M.A. Campanero, and M.J. Blanco-Prieto. 2010. Lipid raft-targeted therapy in multiple myeloma. Oncogene. 29:3748–3757.

Mulloy, J.C., J.A. Cancelas, M.-D. Filippi, T.A. Kalfa, F. Guo, and Y. Zheng. 2010. Rho GTPases in hematopoiesis and hemopathies. Blood. 115:936–947.

Olivo-Marin, J.-C. 2002. Extraction of spots in biological images using multiscale products. Pattern Recogn. 35:1989–1996.

Orkin, S.H., and L.I. Zon. 2008. SnapShot: hematopoiesis. Cell. 132:712.

Palumbo, A., and K. Anderson. 2011. Multiple myeloma. The New England journal of medicine. 364:1046–1060.

Pichiorri, F., S.S. Suh, M. Ladetto, M. Kuehl, T. Palumbo, D. Drandi, C. Taccioli, N. Zanesi, H. Alder, J.P. Hagan, R. Munker, S. Volinia, M. Boccadoro, R. Garzon, A. Palumbo, R.I. Aqeilan, and C.M. Croce. 2008. MicroRNAs regulate critical genes associated with multiple myeloma pathogenesis. Proceedings of the National Academy of Sciences of the United States of America. 105:12885–12890.

Plebanek, M.P., R.K. Mutharasan, O. Volpert, A. Matov, J.C. Gatlin, and C.S. Thaxton. 2015. Nanoparticle Targeting and Cholesterol Flux Through Scavenger Receptor Type B-1 Inhibits Cellular Exosome Uptake. Sci Rep. 5:15724.

Prince, H.M., M. Bishton, and S. Harrison. 2008. The potential of histone deacetylase inhibitors for the treatment of multiple myeloma. Leuk Lymphoma. 49:385–387.

Raje, N., and D.L. Longo. 2015. Monoclonal Antibodies in Multiple Myeloma Come of Age. The New England journal of medicine. 373:1264–1266.

Ren, Z., Y. Su, and X. Liu. 2024. ChatGPT-Powered Hierarchical Comparisons for Image Classification. NIPS ’23: Proceedings of the 37th International Conference on Neural Information Processing Systems:69706–69718.

Roccaro, A.M., A. Sacco, B. Thompson, X. Leleu, A.K. Azab, F. Azab, J. Runnels, X. Jia, H.T. Ngo, M.R. Melhem, C.P. Lin, D. Ribatti, B.J. Rollins, T.E. Witzig, K.C. Anderson, and I.M. Ghobrial. 2009. MicroRNAs 15a and 16 regulate tumor proliferation in multiple myeloma. Blood. 113:6669–6680.

Rosin, P.L. 2001. Unimodal thresholding. Pattern Recognition. 34:2083–2096.

Rozic, G., P. Lena, J. Jakubikova, D. Adrian, A. Avigdor, A. Nagler, and M. Leiba. 2015. STK405759 As a Novel Tubulin Active Agent for Multiple Myeloma Therapy. Blood. 126:5348–5348.

Rublee, E., V. Rabaud, K. Konolige, and G. Bradski. 2011. ORB: an efficient alternative to SIFT or SURF. International Conference on Computer Vision:2564–2571.

Saviana, M., P. Le, L. Micalo, D. Del Valle-Morales, G. Romano, M. Acunzo, H. Li, and P. Nana-Sinkam. 2023. Crosstalk between miRNAs and DNA Methylation in Cancer. 14:1075.

Scovanner, P., S. Ali, and M. Shah. 2007. A 3-dimensional sift descriptor and its application to action recognition. In Proceedings of the 15th ACM international conference on Multimedia. ACM, Augsburg, Germany. 357–360.

Shi, J., and C. Tomasi. 1994. Good features to track. IEEE Conference on Computer Vision and Pattern Recognition:593–600.

Silva, A., M.C. Silva, P. Sudalagunta, A. Distler, T. Jacobson, A. Collins, T. Nguyen, J. Song, D.T. Chen, L. Chen, C. Cubitt, R. Baz, L. Perez, D. Rebatchouk, W. Dalton, J. Greene, R. Gatenby, R. Gillies, E. Sontag, M.B. Meads, and K.H. Shain. 2017. An Ex Vivo Platform for the Prediction of Clinical Response in Multiple Myeloma. Cancer Res. 77:3336–3351.

Spicka, I., E.M. Ocio, H.E. Oakervee, R. Greil, R.H. Banh, S.Y. Huang, J.M. D’Rozario, M.A. Dimopoulos, S. Martinez, S. Extremera, C. Kahatt, V. Alfaro, A.M. Carella, N. Meuleman, R. Hajek, A. Symeonidis, C.K. Min, P. Cannell, H. Ludwig, P. Sonneveld, and M.V. Mateos. 2019. Randomized phase III study (ADMYRE) of plitidepsin in combination with dexamethasone vs. dexamethasone alone in patients with relapsed/refractory multiple myeloma. Ann Hematol. 98:2139–2150.

Sung, M.S., A. Gjyrezi, G.Y. Lee, A. Matov, G. Galletti, M. Loftus, Y. Syed, T. Lannin, A. Hristov, C. Mason, S.T. Tagawa, B.J. Kirby, D.M. Nanus, and P. Giannakakou. 2013. Using CTCs to interrogate mechanisms of taxane resistance in the prospective TAXYNERGY clinical trial in prostate cancer. Cancer Research. 73:3492.

Sung, M.S., A. Gjyrezi, Y. Syed, T. Lannin, A. Hristov, A. Matov, M. Loftus, G. Galletti, G.Y. Lee, C. Mason, B.J. Kirby, D.M. Nanus, S.T. Tagawa, and P. Giannakakou. 2012. Molecular determinants of taxane activity using circulating tumor cells (CTCs) from castrate-resistant prostate cancer patients. Understanding Cancer: Genomes to Devices Symposium, New York Academy of Medicine.

Suzuki, H., R. Maruyama, E. Yamamoto, and M. Kai. 2012. DNA methylation and microRNA dysregulation in cancer. Mol Oncol. 6:567–578.

Tasaki, S., M. Sung, A. Matov, G. Galletti, E. Diamond, N. Bander, K. Zhou, S. Tagawa, D. Nanus, and P. Giannakakou. 2014. Analysis of microtubule perturbations and androgen receptor localization in circulating tumor cells from castration resistant prostate cancer patients as predictive biomarkers of clinical response to docetaxel chemotherapy. Cancer Research. 74:923–923.

Thery, C., K.W. Witwer, E. Aikawa, M.J. Alcaraz, J.D. Anderson, R. Andriantsitohaina, A. Antoniou, T. Arab, F. Archer, G.K. Atkin-Smith, D.C. Ayre, J.M. Bach, D. Bachurski, H. Baharvand, L. Balaj, S. Baldacchino, N.N. Bauer, A.A. Baxter, M. Bebawy, C. Beckham, A. Bedina Zavec, A. Benmoussa, A.C. Berardi, P. Bergese, E. Bielska, C. Blenkiron, S. Bobis-Wozowicz, E. Boilard, W. Boireau, A. Bongiovanni, F.E. Borras, S. Bosch, C.M. Boulanger, X. Breakefield, A.M. Breglio, M.A. Brennan, D.R. Brigstock, A. Brisson, M.L. Broekman, J.F. Bromberg, P. Bryl-Gorecka, S. Buch, A.H. Buck, D. Burger, S. Busatto, D. Buschmann, B. Bussolati, E.I. Buzas, J.B. Byrd, G. Camussi, D.R. Carter, S. Caruso, L.W. Chamley, Y.T. Chang, C. Chen, S. Chen, L. Cheng, A.R. Chin, A. Clayton, S.P. Clerici, A. Cocks, E. Cocucci, R.J. Coffey, A. Cordeiro-da-Silva, Y. Couch, F.A. Coumans, B. Coyle, R. Crescitelli, M.F. Criado, C. D’Souza-Schorey, S. Das, A. Datta Chaudhuri, P. de Candia, E.F. De Santana, O. De Wever, H.A. Del Portillo, T. Demaret, S. Deville, A. Devitt, B. Dhondt, D. Di Vizio, L.C. Dieterich, V. Dolo, A.P. Dominguez Rubio, M. Dominici, M.R. Dourado, T.A. Driedonks, F.V. Duarte, H.M. Duncan, R.M. Eichenberger, K. Ekstrom, S. El Andaloussi, C. Elie-Caille, U. Erdbrugger, J.M. Falcon-Perez, F. Fatima, J.E. Fish, M. Flores-Bellver, A. Forsonits, A. Frelet-Barrand, et al. 2018. Minimal information for studies of extracellular vesicles 2018 (MISEV2018): a position statement of the International Society for Extracellular Vesicles and update of the MISEV2014 guidelines. Journal of extracellular vesicles. 7:1535750.

van de Wetering, M., H.E. Francies, J.M. Francis, G. Bounova, F. Iorio, A. Pronk, W. van Houdt, J. van Gorp, A. Taylor-Weiner, L. Kester, A. McLaren-Douglas, J. Blokker, S. Jaksani, S. Bartfeld, R. Volckman, P. van Sluis, V.S. Li, S. Seepo, C. Sekhar Pedamallu, K. Cibulskis, S.L. Carter, A. McKenna, M.S. Lawrence, L. Lichtenstein, C. Stewart, J. Koster, R. Versteeg, A. van Oudenaarden, J. Saez-Rodriguez, R.G. Vries, G. Getz, L. Wessels, M.R. Stratton, U. McDermott, M. Meyerson, M.J. Garnett, and H. Clevers. 2015. Prospective derivation of a living organoid biobank of colorectal cancer patients. Cell. 161:933–945.

Vaswani, A., N. Shazeer, N. Parmar, J. Uszkoreit, L. Jones, A.N. Gomez, Ł. Kaiser, and I. Polosukhin. 2017. Attention is all you need. In Proceedings of the 31st International Conference on Neural Information Processing Systems. Curran Associates Inc., Long Beach, California, USA. 6000–6010.

Wang, J., A. Hendrix, S. Hernot, M. Lemaire, E. De Bruyne, E. Van Valckenborgh, T. Lahoutte, O. De Wever, K. Vanderkerken, and E. Menu. 2014. Bone marrow stromal cell-derived exosomes as communicators in drug resistance in multiple myeloma cells. Blood. 124:555–566.

Wang, S.Y., T. Holzhey, S. Heyn, T. Zehrfeld, S. Fricke, F.A. Hoffmann, C. Becker, L. Braunert, T. Edelmann, I. Paulenz, M. Hitzschke, F. Flade, A. Schwarzer, K. Fenchel, G.N. Franke, V. Vucinic, M. Jentzsch, S. Schwind, S. Hell, D. Backhaus, T. Lange, D. Niederwieser, M. Scholz, U. Platzbecker, and W. Pönisch. 2023. Impact of the changing landscape of induction therapy prior to autologous stem cell transplantation in 540 newly diagnosed myeloma patients: a retrospective real-world study. Journal of cancer research and clinical oncology. 149:3739–3752.

Zhang, W., Y. Gu, Q. Sun, D.S. Siegel, P. Tolias, Z. Yang, W.Y. Lee, and J. Zilberberg. 2015. Ex Vivo Maintenance of Primary Human Multiple Myeloma Cells through the Optimization of the Osteoblastic Niche. PloS one. 10:e0125995.

Zhang, W., W.Y. Lee, D.S. Siegel, P. Tolias, and J. Zilberberg. 2014. Patient-specific 3D microfluidic tissue model for multiple myeloma. Tissue Eng Part C Methods. 20:663–670.

Zheng, Y., C. Tu, J. Zhang, and J. Wang. 2019. Inhibition of multiple myelomaderived exosomes uptake suppresses the functional response in bone marrow stromal cell. International journal of oncology. 54:1061–1070.

